# The impact of self-isolation on psychological wellbeing and how to reduce it: a systematic review

**DOI:** 10.1101/2023.10.16.23296895

**Authors:** Alex F. Martin, Louise E. Smith, Samantha K. Brooks, Madeline V. Stein, Rachel Davies, Richard Amlôt, Neil Greenberg, G James Rubin

**Affiliations:** King’s College London, Institute of Psychiatry, Psychology and Neuroscience, London, UK; NIHR Health Protection Research Unit in Emergency Preparedness and Response, London, UK; UK Health Security Agency, Chief Scientific Officer’s Group, UK

**Keywords:** mental health, COVID-19, public health, quarantine, isolation, factors associated, narrative synthesis

## Abstract

Self-isolation is a public health measure used to prevent the spread of infection, and which can have an impact on the psychological wellbeing of those going through it. It is likely that self-isolation will be used to contain future outbreaks of infectious disease. We synthesised evidence on the impact of home self-isolation on psychological wellbeing of the general public during the COVID-19 pandemic.

This systematic review was registered on PROSPERO (CRD42022378140). We searched Medline, PsycINFO, Web of Science, Embase, and grey literature (01 January 2020 to 13 December 2022). Our definition of wellbeing included adverse mental health outcomes and adaptive wellbeing. Studies that investigated isolation in managed facilities, children, and healthcare workers were excluded. We followed PRISMA and synthesis without meta-analysis (SWiM) guidelines. We extracted data on the impact of self-isolation on wellbeing, and factors associated with and interventions targeting wellbeing during self-isolation.

We included 36 studies (most were cross sectional, two were longitudinal cohort studies, three assessed interventions, and five were qualitative). The mode quality rating was ‘high-risk’. Depressive and anxiety symptoms were most investigated. Evidence for an impact of self-isolation on wellbeing was often inconsistent in quantitative studies, although qualitative studies consistently reported a negative impact on wellbeing. However, people with pre-existing mental and physical health needs consistently reported increased symptoms of mental ill health during self-isolation. Studies reported modifiable stressors that have been reported in previous infectious disease contexts, such as inadequate support, poor coping strategies, inadequate and conflicting information, and the importance of regular contact from trusted healthcare professionals. However, interventions targeting psychological wellbeing were rare and evaluative studies of these had high or very high risk of bias.

When implementing self-isolation directives, public health officials should prioritise support for more vulnerable individuals who have pre-existing mental or physical health needs, lack support, or who are facing significant life stressors. Clinicians can play a key role in identifying and supporting those most at risk. Focus should be directed toward interventions that address loneliness, worries, and misinformation, whilst monitoring and identifying individuals in need of additional support.

## Introduction

To minimise the spread of infection and protect the public, several containment measures have been used in infectious disease outbreaks, including isolation (the separation of those who are ill from those who are well) and quarantine (the separation of those at risk of developing an illness from those who are well). During the COVID-19 pandemic, there was a substantial focus among public health practitioners across the world on the use of these strategies (hereafter *self-isolation*) ^1^

Self-isolation can be a time of upheaval and concern with potential impacts on the psychological wellbeing of those going through it. For example, isolation may lead to fears about illness progression, or worries about financial loss and caretaking responsibilities. Psychological wellbeing (hereafter *wellbeing*) encompasses both adverse mental health outcomes, such as symptoms and disorders, and adaptive outcomes, such as resilience and flourishing. In February 2020, Brooks and colleagues published a rapid review of studies that assessed the psychological impact of quarantine.^2^ The findings indicated that quarantine was associated with adverse wellbeing outcomes in nearly all contexts, with some evidence that these effects could be long-lasting. A subsequent meta-analysis found that the odds of depressive, anxiety, and stress related disorders was more than double for people who self-isolated compared to those who had not.^3^ The study also reported an increased risk for some broader wellbeing outcomes, such as insomnia and substance use. All the studies included in these reviews were conducted before the COVID-19 pandemic.

The COVID-19 pandemic led to a proliferation of studies on wellbeing and its associated factors. For example, pre-existing mental health disorders, loneliness, worry, and a lack of access to resources and support were all identified as risk factors for poorer mental health during the pandemic.^4–6^ Conversely, an editorial published during the early stages of the pandemic reported that isolation could have a positive ‘downstream’ impact on wellbeing through, for example, increased health behaviours such as physical activity and healthy eating.^7^ During the pandemic, many studies investigated the impact on wellbeing of ‘lockdown’ measures. Lockdown, in contrast to self-isolation, involved population-wide ‘stay-at-home’ or ‘mass quarantine’ orders, where people were required to stay at home except for essential activities and exercise. What is less clear is how self-isolation specifically impacted wellbeing, given that this containment measure has the potential for a greater impact on specific aspects of mental health, such as social stigma and anxiety related to the likelihood of infection or the development of symptoms.^2,3^

A recent review of 25 studies carried out globally found that testing followed by self-isolation was an important public health mitigation measure to reduce transmission during the COVID-19 pandemic.^8^ It is likely that home-based self-isolation will be used in future outbreaks of infectious disease, as it was during the UK public health response to the 2022 mpox outbreak.^9^ Reducing the burden of self-isolation on those affected remains a scientific and policy priority.

This systematic review appraises:

1. The impact of self-isolation on wellbeing during or following the COVID-19 pandemic.
2. Factors associated with wellbeing outcomes during or following self-isolation.
3. The effectiveness of interventions designed to improve wellbeing during or following self-isolation.

## Methods

This systematic literature review was carried out in accordance with the Cochrane Collaboration guidelines for the conduct of systematic reviews,^10^ and the Preferred Reporting Items for Systematic Reviews and Meta-Analyses (PRISMA; see Appendix 1).^11,12^ The protocol was prospectively registered on PROSPERO (CRD42022378140).

### Search strategy and selection criteria

A systematic search was conducted of studies published between 01 January 2020 and 13 December 2022. We searched six databases (Medline, PsycInfo, Web of Science, Embase, PsyArXiv, medRxiv). The search strategy included terms for COVID-19, isolation and quarantine (combined with NOT social isolation), and wellbeing. The search was also used for a separate systematic review exploring adherence to self-isolation,^13^ screening was performed in parallel up to full-text screening. We searched five grey literature databases, relevant UK agencies and organisations, Google, and made direct inquiries with UK Government agencies. A full description of the search is reported in Appendix 2.

The search was piloted, and the reviewing team (AFM, LES, SKB, MVS, RD, and GJR) reviewed a training set of 300 studies. Discrepancies were discussed until agreement on included studies was attained. Piloting led to some revisions and clarifications of the protocol (Appendix 3). Then, reviewers independently screened citations, meeting weekly to reach agreement on queries and discrepancies.

Studies were included if they used original data to investigate the impact of self-isolation on wellbeing during the COVID-19 pandemic. Self-isolation was defined as: anyone advised (directly or by widely disseminated public health guidance) to avoid contact with others because they were known or suspected to have COVID-19 or because they were suspected to be incubating COVID-19. Wellbeing was broadly defined, including adverse mental health outcomes and adaptive characteristics. We included adults who self-isolated at home and excluded children, healthcare workers, and those in managed isolation facilities or in a hospital. For aim 1, quantitative studies had to use a design that allowed attribution of the impact of self-isolation on wellbeing, for example, through use of a comparison to a control group. For aim 2, factors associated with wellbeing had to be directly related to, or occur during, the self-isolation period. For example, studies investigating the impact of a change in the national containment rules after the isolation period but before the study was carried out were excluded. For this aim, studies were included that compared home to isolation in a managed facility. Grey literature was only included if it investigated the effectiveness of an intervention, to ensure only the most rigorous non-peer-reviewed studies were included and because of the dearth of peer-reviewed data on this specific topic. If it was unclear whether a study met the inclusion criteria, the corresponding author was contacted, and the study was excluded if no response was received.

### Data Analysis

Data were extracted by one reviewer (AFM for quantitative studies and SKB for qualitative studies) using a piloted, standardised table. All studies were discussed with at least one other reviewer (AFM, LES or GJR). Extracted data included: study characteristics (design, methods, sampling, demographics); isolation characteristics (reason, duration, context); and wellbeing characteristics (measures, impact, associated factors, interventions). We reported the most rigorous analysis conducted in each study, for example, multivariable analysis over unadjusted analysis.

Study quality assessment was performed by one reviewer (AFM for quantitative studies and SKB for qualitative studies), all studies were discussed with at least one other reviewer (AFM, LES or GJR). For quantitative studies, we used the Risk of Bias in Non-randomized Studies for Exposure (ROBINS-E) and Interventions (ROBINS-I).^14,15^ ROBINS assessments are specific to a reported result rather than a study. Consequently, studies that reported a result for aim 1 and 2 received two risk of bias scores. Each result was categorised as low risk, some concerns, high risk, or very high risk based on the tool’s algorithm.

We used the Critical Appraisal Skills Programme (CASP) checklist for qualitative studies,^16^ but reworded the item ‘how valuable is the research?’ to ‘do the authors discuss the value of the research in terms of implications and contribution to literature?’ to allow yes/no responses in line with the other items and to give each study an overall quality score percentage. Scores were out of ten, reported as a percentage, with higher scores indicating better quality. Risk of bias tools are reported in Appendix 4.

Quantitative data were synthesised narratively, following SWiM guidelines.^17^ No meta-analysis was planned due to expected heterogeneity in study design, outcomes, and associated factors. Qualitative data were synthesised using meta-ethnography, following eMERGe guidelines.^18^ A description of the synthesis of results for each aim is reported in Appendix 5.

## Results

The search identified 15,275 citations (Figure 1). Thirty-six studies were included, all of which were identified through database searches.^19–54^

**Figure 1.**
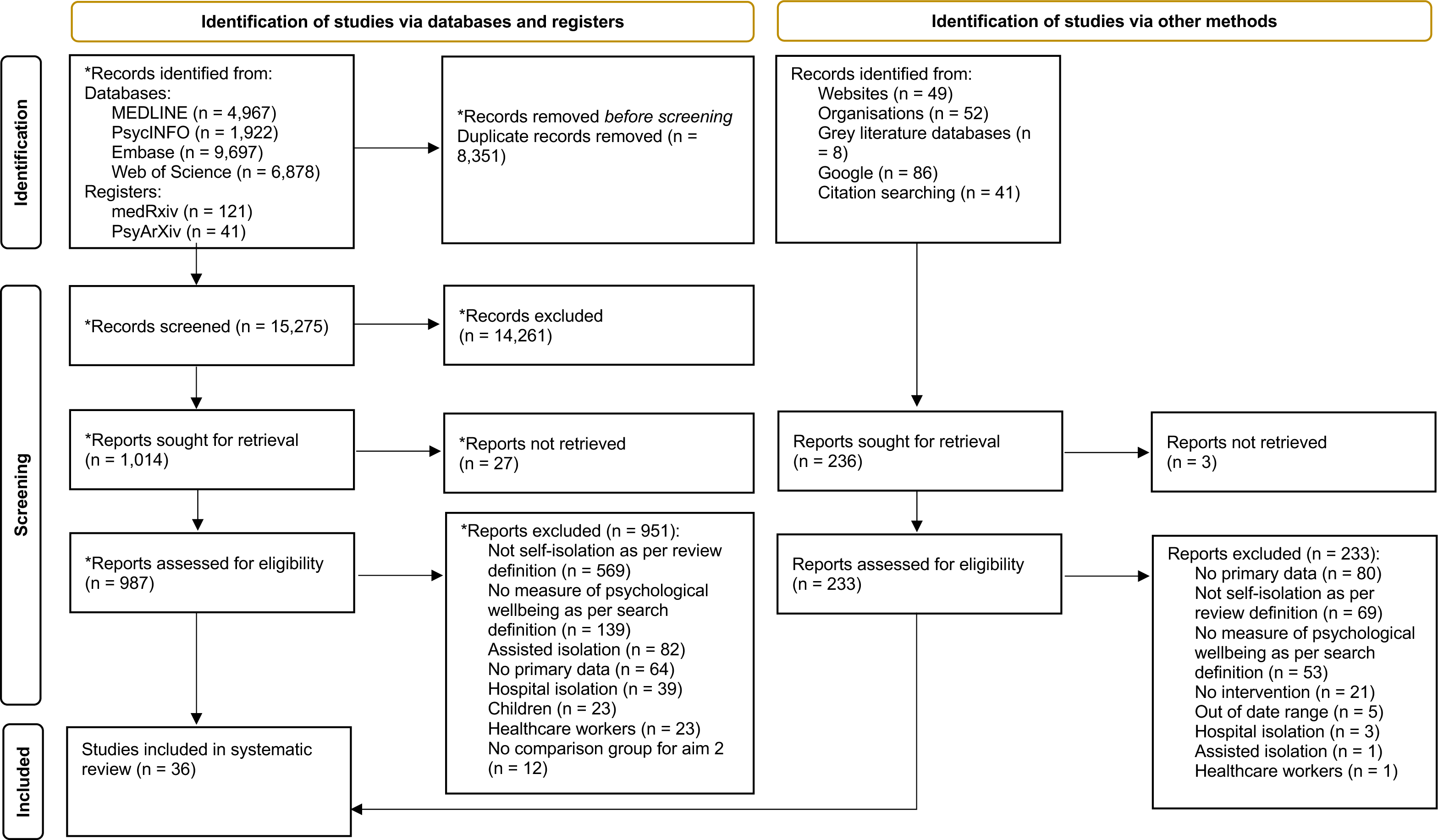
Study selection flowchart. <u>Note. *</u>At this stage, citation screening was completed for this systematic review and a systematic review investigating adherence to self-isolation. Therefore, these totals include citations screened both systematic reviews.

### Study characteristics

Most of the studies were conducted in Europe,^19,23,26,30,34,36–39,41,45,47,50,52,53^ Asia,^22,24,28,29,33^ and South Asia.^20,31,46,48,49^ The rest were in Africa,^21,40,42,43^ East Asia,^32,35,54^ South America,^27,44^ North America,^25^ and one study was multi-continent.^51^ Most studies were cross-sectional in design,^19–23,25,27,29,30,32,34,36,37,39–45,47,49–51,53,54^ two were longitudinal,^35,46^ three were interventions,^24,31,48^ and five were qualitative.^26,28,33,38,52^ Sample sizes ranged from 14 to 18,146. One study included only older adults,^22^ all other studies included adults aged 18 years and over.

Studies often reported on more than one wellbeing outcome. The most reported outcomes were anxiety symptoms,^19,24,27,31,35,36,39,40,42–45,47,48,50,51,54^ depressive symptoms,^19,27,31,32,35,36,39,40,42–45,48–51^ and general psychological symptoms.^19,20,25,27,30,31,34,37,39,43,46,48^ Only three quantitative studies reported adaptive wellbeing outcomes.^31,46,48^ Reasons for self-isolation were COVID-19 infection,^21,24,26,28,29,31,33,35,37,40,41,43,45,47–49^ suspected infection,^42^ close contact with an infected person,^30,36,39,52,54^ or a combination of these.^19,20,22,23,25,27,32,34,38,44,50,51,53^ One study did not report a reason.^46^

Ten studies were carried out during national/regional lockdown measures,^20,23,27,29,41,42,45,46,49,50^ two were not,^25,39^ two were mixed,^36,51^. Twenty-two studies did not report lockdown context, all but one^19^ related to aims 2 and 3.^21,22,24,26,28,30–35,37,38,40,43,44,47,48,52–54^

Study characteristics are reported in Table 1.

**Table 1.** Study characteristics.

|  | Country; dates of data collection | Study design; data collection method. | Sample frame; sampling method; sample size; response rate | Study inclusion criteria | Percent female; age: mean (standard deviation), unless otherwise stated; group size (percentage) |
| --- | --- | --- | --- | --- | --- |
| **Aaltonen et al., 2022** <sup>19</sup> | Finland; 12 May to 23 June 2020. | Longitudinal (aim 1), cross sectional (aim 2); telephone interview. | Population based; identified from the register of the infectious diseases control unit in the city of Kerava using random sampling; N=112; 98%. | ≥18 years old; Finnish-speaking persons; suspected infection between 19 May and 25 June 2020. | Quarantine group N=43<br>58%; 40.9y (12.1).<br>Self-isolation group N=14<br>36.1y (11.2).<br>Control group (negative PCR, N=523) 58%; 43.5y (14.0). |
| **Abir et al., 2021** <sup>20</sup> | Bangladesh; 1 to 30 April 2020. | Cross sectional; online survey. | Population based; disseminated through social media and snowball sampling; N=10,609; not reported. | ≥18 years old; resident of Bangladesh. | 50.4%; The majority (57.0%) of respondents were young adults aged 18 to 27y; na. |
| **Aloba & Opakunle, 2021** <sup>21</sup> | Nigeria; October 2020 to January 2021. | Cross sectional; online survey. | Population based; participants presented to one of two treatment centres in the capital of Nigeria; 'eligible Nigerian adults with COVID-19 were recruited'; N=498; not reported. | ≥18 years old; stable internet access required; positive PCR.<br>Exclusion criteria: pre-existing mental health disorders or co-morbid medical disorders | 47.3%; 41.3y (14.6), range=18-80; na. |
| **Aslaner et al., 2022** <sup>22</sup> | Turkey; 30 September 2020 to 10 January 2021. | Cross sectional; telephone interview. | Older adults; identified from local government records in Kayseri; N=656 included; 81% of those contacted. | ≥65 years old. | 51.5%; 69.9y (5.7), range=65-80; na. |
| **Bonsaksen et al., 2020** <sup>23</sup> | Norway; 8 April to 20 May 2020. | Cross sectional; online survey. | Population based; identified from the Norwegian cross-sectional survey CORONAPOP, which collected data by means of an open web-link disseminated from several institutions, including Oslo hospitals, social media, and national and local newspapers; N=4527; not reported. | ≥18 years old; Norwegian citizens | 85.4% (of the 4,509 who reported data on gender);<br>18–29y: N=1156 (25.5%)<br>30–39y: N=1220 (26.9%)<br>40–49y: N=931 (20.6%)<br>50–59y: N=766 (16.9%)<br>60–69y: N=354 (7.8%)<br>70y+: N=100 (2.2%); na. |
| **Chakeri et al., 2020** <sup>24</sup> | Iran; not reported. | Intervention – telenursing counselling every other day for 3 weeks. | Population based; recruited through emergency departments of two hospitals using continuous sampling; N=100; not reported. | Patients who presented to the emergency department and were given a diagnosis of COVID-19 after a CT scan of their lungs; the physician prescribed home quarantine, medication, and continued treatment at home. | Control group (N=50)<br>Not reported; 42.4y (9.0)<br>Intervention group (N=50)<br>Not reported; 42.7y (9.4). |
| **Daly et al., 2021**<br><sup>25</sup> | Canada; 14 to 29 May 2020. | Cross sectional; online survey. | Population based; survey distributed by a national polling vendor, the total panel includes 125,000 adults from all provinces and territories. Increased recruitment of panel participants from traditionally under-represented groups. Participants were randomly sampled using stratification of sociodemographic characteristics to ensure representative of Canadian population with adjustments for response propensity; 3558 were invited to reach 3000 (84% response rate). | ≥18 years old; resident in Canada. | 50.6%;<br>18-34y: N=534 (17.8%)<br>35-54y: N=1157 (38.6%)<br>55y+: N=1309 (43.6%); na. |
| **Domenghino et al., 2022** <sup>26</sup> | Switzerland; participants enrolled 6 October 2020 to 26 January 2021 (retrospective group) and 6 August 2020 to 19 January 2021 (prospective group). | Qualitative*; free text survey responses.<br>*\*only the qualitative findings met the review inclusion criteria* | Population based; identified from the Zurich SARS-CoV-2 cohort study, individuals with SARS-CoV-2 infection reported to authorities. Two populations were recruited, the first 'retrospectively recruited' (infected prior to the study) and the second 'prospectively recruited', an age-stratified random sample of all eligible individuals; N=1,547; For the retrospectively recruited population: N=442; 33.7%, for the prospectively recruited population: N=1,105; 34.5%. | ≥18 years old; residing in the Canton of Zurich; able to follow study procedures; sufficient knowledge of German language; diagnosed with SARS-CoV-2 infection either before the study (between 27 February 2020 and 05 August 2020, retrospective group) or between 06 August 2020 and 19 January 2021 (prospective group). | 50.6%; 49.2y (range 17-92)*; na.<br>*\*being 18 or over was one of the inclusion criteria; unclear why 17-year-olds were included* |
| **Flores-Torres et al., 2021** <sup>27</sup> | Mexico; 4 June to 8 July 2020. | Cross sectional; online survey. | Population based; identified from the Study of Urban Health and Social Distancing (SUSana) of Mexico City government employees through an email invite N=2,016; 14.0%. | All employees with access to an institutional email<br>Exclusion criteria: employees from Mexico City's Ministry of Health. | 49.8%; 42.6y (12.1); na. |
| **Gok, 2022** <sup>28</sup> | Turkey; 'Procedure' subsection of the Methods states data was collected between 1 April to 30 May 2021, however, the 'Sample' subsection of the Methods states that data collection started 'in March 2021 and was completed in about a month' - unclear which of these is correct. | Qualitative; free text survey responses. | Population based; individuals diagnosed with COVID-19 living in Turkey, recruited via 'appropriate sampling' which is the sample of participants who can be 'accessed more effortlessly because of limitations such as time and place'; N=212; not reported. | Living in Turkey; diagnosis of COVID-19. | 72.2%; 35.8y (9.67)<br>Range 18-60<br>18-25: N=37 (17.4%)<br>26-35: N=71 (33.4%)<br>36-46: N=73 (34.4%)<br>47-60: N=31 (14.8%), na. |
| **Havlioglu et al., 2022** <sup>29</sup> | Turkey; January – April 2021. | Cross sectional; online questionnaire. | Population based; all individuals in region with a positive PCR; N=800; 28.2%. | ≥18-year-old; in the Sanliurfa province; positive PCR. | 45.1%; 18-30 N=539 (44.1%) no other age data reported; na. |
| **Isherwood et al., 2022** <sup>30</sup> | Wales; Round 1, 11 November to 1 December 2020 and Round 2, 18 February to 23 March 2021. | Cross sectional; telephone interview. | Population based; all individuals who were a close contact of a confirmed case, quota sampling based on age, gender, and SES; N=2,027; 18.8%. | ≥18 years old; resident in Wales; successfully contacted by TTP after forward contact tracing; completed their self-isolation period at the time of telephone survey.<br>Exclusion criteria: currently self-isolating; a contact of a case of COVID-19 who died. | 53.6%;<br>18-29: N=598 (29.5%)<br>30-39: N=374 (18.5%)<br>40-49: N=344 (17.0%)<br>50-59: N=409 (20.0%)<br>60+ N=301 (9.6%); na. |
| **Jagadeesan et al., 2022** <sup>31</sup> | India; May to June 2021. | Intervention – Bhramari Pranayama (yogic breathing) 20 mins twice daily (online) for 15 days. | Population based; patients registered in a COVID-19 home care programme; N=42 participated; 70.0%. | 18-60 years old; under home care program of the host institute; met the diagnostic criteria of COVID-19 and asymptomatic; with an internet facility; able to independently cooperate with doctors in online programme.<br>Exclusion criteria: moderate to critical COVID-19 symptoms; pre-existing acute respiratory diseases, asthmatic, heart disease, cognitive impairments, and pregnant women. | 30.0%; 39.2y (14.6); na. |
| **Jang et al., 2022** <sup>32</sup> | South Korea; 16 August to 31 October 2020. | Cross sectional; in person interview. | Population based; identified from the 2020 Korean Community Health Survey (KCHS) using the population stratified sampling from the original study; N=1,071 included; not reported. | ≥19 years old; all individuals in KCHS study who had self-isolated. | 46.6%; 40.0y (standard error=0.31); na. |
| **Jesmi et al., 2021** <sup>33</sup> | Iran; April 2020 to July 2020. | Qualitative; in-person and telephone interviews. | Population based; recruited from Sabzevar Vasei Hospital (Iran) after discharge using purposive and snowball sampling; N=14; not reported. | ≥18 years old; confirmed diagnosis of COVID-19 by a chest CT scan and PCR test; able to communicate and willing to share their experiences | 64%; 37.7y (9.5*); na.<br>*\*calculated by the review authors – the paper only presented individual ages and a range of 25-53* |
| **Joisten et al., 2022** <sup>34</sup> | Germany; Not reported | Cross-sectional, online survey | Population based; taken from the CoCo-Fakt cohort study, identified through government register of infected people and their close contacts*; N=10,490; 31.1%.<br>*\*data from the same study as Wessely et al. (2022), but reporting different outcomes.* | >16 years of age*<br>Exclusion criteria: without an email address, hospitalised or deceased affected, nursing home residents.<br>*\*16 and 17 year olds were included, although the number was not reported, the mean age and SD suggests that they were a small minority and the study was therefore included* | Whole sample: 60.0%, 40.8y (14.2);<br>Infected N=4,065<br>Contacts N=6,425. |
| **Ju et al., 2021** <sup>35</sup> | China; 10 February to 2 April 2020. | Longitudinal, prospective; online questionnaire. | Population based; recruited from The First Hospital of Changsha (Hunan, China) after discharge; N=146 included at baseline (79.8%) | ≥18 years old (one adolescent aged 15 was included); able to use mobile devices to complete the questionnaires. | Whole sample: 46.3%; median = 39y (IQ range 30-47); of 95 included in the study (that completed both timepoints)<br>Home isolation N=45<br>Hotel isolation N=50. |
| **Kopilas et al., 2021** <sup>36</sup> | Croatia and Italy; 4 to 24 March 2020 | Cross sectional*; online questionnaire. | Population based; convenience sampling of universities and colleagues; N=164; 71.3%. | ≥18 years old; speak English. | Whole sample: 69.3%; 37.3y (SD=13.6);<br>Exposed (isolation following close contact) N=27. |
|  |  | *\*only the quantitative findings met the review inclusion criteria* |  |  | Not exposed N=136 (made up of three groups: in lockdown (N=72), no contact (N=21) and unrelated (N=43)). |
| **Kowalski et al., 2021** <sup>37</sup> | Germany; 29 January to 12 April. | Cross sectional; online survey. | Population based; taken from government register of positive PCR cases; N=224 included; 37.5%. | ≥18-year-old; within the jurisdiction of the Freudenstadt Health Department; all individuals with a positive PCR.<br>Exclusion criteria: in inpatient care facilities; language barrier to questionnaire completion. | Whole sample: 52.7%; 41y (14.6); With psychological burden, N=104 Without psychological burden N=121. |
| **Lohiniva et al., 2021** <sup>38</sup> | Finland; April to May 2020. | Qualitative; in-person interviews. | Population based; participants were PCR confirmed cases of COVID-19 (and their families) recruited via the Finnish Institute for Health and Welfare website or an SMS sent to confirmed cases, sampling was based on 'maximum variation' to engage participants from different types of households – this study is part of a larger study on COVID-19 transmission where individuals recruited for that study could also take part in this qualitative study if they wished; N=64 participants from 24 households; not reported. | ≥12 years old*; households located in Helsinki; households with at least one COVID-19 PCR confirmed case and at least one additional person in the household, 'in home quarantine or in isolation for a period of time'.<br>*\*only those participants 18+ have had their data extracted due to this review's inclusion criteria* | Of the 50 adults: gender % not reported ('the sample included approximately an equal number of female and male respondents'); mean age not reported, 'most adult participants were in the age range of 30-49 (75%)'; na. |
| **Maric et al., 2022** <sup>39</sup> | Serbia, June to October 2021. | Cross sectional; in-person interviews. | Population based; identified from the Serbian CoV2Soul.rs study using multistage probabilistic sampling from 135 randomly selected local communities in 60 out of the 180 municipalities in Serbia and quasi-randomisation of households in identified areas; N=1,203, 67.0%. | 18-65 years old; resident in identified households; spoke Serbian. | 51.3%; 43.7y (13.6); na. |
| **Mohamed & Yousef, 2021** <sup>40</sup> | Egypt, 22 May to 28 July 2020. | Cross sectional; online survey. | Population based. Patients were recruited from Zagazig University Hospital; N=89; not reported. | ≥18 years old; PCR confirmed mild and moderate cases of COVID-19.<br>Exclusion criteria: Presence of 'mental retardation, dementia, or delirium'; confirmed severe cases of COVID-19. | Home isolated N=43; 33.6%; 39.9y (8.8) Hospital isolated (N=46); 30.4%; 41.3y (9.3). |
| **Navas et al., 2022**<sup>41</sup> | Spain, April to June 2020. | Cross sectional; telephone interview. | Population based; selected from primary care electronic medical records lists from the Passeig de Sant Joan Primary Care Team in Barcelona; N=89; not reported. | ≥18 years old; PCR confirmed infection; have lived with other people in the preceding two weeks.<br>To be eligible to stay in the supervised hotel, they had to fulfil at least one of the following criteria: caring for a vulnerable person; being destitute or homeless; being a tourist in transit; sharing their home with many people; fulfilling other vulnerability requirements as assessed by a social worker. | Whole sample: 64%; 53.6y (16.9); Home isolated N=45 Hotel isolated N=44. |
| **Oginni et al., 2021**<sup>42</sup> | Nigeria; not reported. | Cross sectional; online survey. | Population based; recruited through social media and WhatsApp, participants represented all Nigerian tribes; N=966; not reported. | ≥18 years old; resident in Nigeria for at least six months prior to the lockdown; fluent in English; able to use the internet;<br>Exclusion criteria: severe cognitive or physical impairments. | 49.6%; median = 27y (IQR=12y); na. |
| **Opakunle, 2022**<sup>43</sup> | Nigeria; October 2020 to February 2021. | Cross-sectional; online survey. | Population based; convenience sampling via email and WhatsApp; N=509; not reported. | 18 to 80 years old; no pre-existing mental disorder or co-morbid medical problems.<br>Exclusion criteria: too physically ill during isolation; admitted to hospital or isolation centre; given oxygen therapy. | 47.3%; 41.2y (14.6); na. |
| **Paz et al., 2020**<sup>44</sup> | Ecuador; 22 March to 18 April 2020. | Cross-sectional; online survey. | Population based; taken from the surveillance program for COVID-19 established by the Ecuadorian Ministry of Public Health, a non-probabilistic sampling strategy was used from the beginning of the study until quota was reached; N=759; 88.7% response rate. | ≥18 years old; living in Ecuador; under surveillance for diagnosis or suspected illness. | 51.9%; 37.7y (11.0) range 18-94; na. |
| **Petrocchi et al., 2021** <sup>45</sup> | Switzerland; COVID-19 positive (groups 1&2), 4 August to 5 October 2020. COVID-19 negative (group 3), 8 September to 15 October 2020. | Cross sectional; online survey or paper survey (optional). | Population based; random sampling to produce several groups with different sampling methods:<br>COVID-19 positive groups: hospital database of COVID-19 cases<br>1. Home isolated; N=63; 12.6%<br>2. Hospital isolated; N=76; 24.5%<br>3. COVID-19 negative group: university Facebook advert<br>N=61; 31.3%. | ≥18 years old. | Home isolated group N=63<br>60.3%; 41.9y (14.3) range=19–75<br>Hospital isolated group N=76<br>26.3%; 62.1y (14.8) range=24–87<br>Covid negative group N=61<br>83.6%; 41.3y (13.6) range=19–74. |
| **Pheh et al., 2020** <sup>46</sup> | Malaysia; not reported. | Intervention*; online survey<br>*\*this study reports an RCT during nationwide stay at home orders. Here, data is reported related to aim 1 and data is used as a longitudinal, prospective design.* | Population based; convenience and snowball sampling using authors' social media; N=161; 68.5%. | Not reported. | 73.9%; 28.8y (9.0) range=18-70;<br>Experienced self-isolation N=16 (9.9%) |
| **Plesea-Condratovici et al., 2022** <sup>47</sup> | Romania; not reported. | Cross sectional; telephone interview. | Population based; recruited from GP monitoring of COVID-19 positive patients from two GP settings; N=107; not reported. | Not reported. | 42%; 45.1y (15.2) range 13-83; na. |
| **Rajagopalan et al., 2022** <sup>48</sup> | India; not reported. | Intervention: OM chanting and meditation 20 minutes twice a day for 14 days. | Population based; recruited from the home care program of Saveetha medical college and hospital; N=25 included; 28.4%. | 18-60 years old; asymptomatic COVID-19 patients with mild severity.<br>Exclusion criteria: pregnant women; moderate or severe symptoms; other comorbid conditions. | 40.0%; range 41-60 years; na. |
| **Ripon et al., 2020** <sup>49</sup> | Bangladesh; 10 to 29 May 2020. | Cross sectional; online survey. | Population based; social media; N=579; not reported. | ≥18 years old, Bangladeshi citizens; diagnosis by PCR.<br>Exclusion criteria: did not complete the 10th grade or below of education; without a job. | Not reported;<br>18-30y: 17.3%<br>31-45y: 32.5%<br>46-55y: 30.1%<br>>55y: 19.8%.<br>Home quarantine N=3952 (68.2%)<br>Institutional quarantine N=1840 (31.8%) |
| **Rossi et al., 2020**<sup>50</sup> | Italy; 27 March to 6 April 2020. | Cross sectional; online survey. | Population based; paid social media advert plus snowballing; N=18,146; not reported. | ≥18 years old; Italian citizens. | 79.5%; median = 38y (IQR=23y); C Currently in quarantine N=141 (0.8%). |
| **Schluter et al., 2022**<sup>51</sup> | 8 countries and territories: Canada, USA, England, Switzerland, Belgium, Philippines, New Zealand and Hong Kong; 6 to 18 November 2020. | Cross sectional; online survey. | Population based; representative quota sampling for each country (age, gender, region), collected by two polling forms using telephone, online and offline recruitment; N=9027; not reported. | ≥18 years old; resident in one of eight included countries. | 52% (0.5% did not identify with male or female); 47.0y (17.0) range=18-99; No isolation N=5753<br>COVID contact N=566<br>COVID symptoms N=720<br>COVID diagnosis N=457<br>Travel/health N=1199. |
| **Verberk et al., 2021**<sup>52</sup> | Netherlands (N=10) and Belgium (N=8); 5 May to 9 July 2020. | Qualitative*; telephone interview.<br>*\*only the qualitative findings met the review inclusion criteria* | Population based; for the overall study, patients with confirmed positive tests in the Netherlands or Belgium - identified via drive-through testing sites, healthcare worker screening, hospital emergency visits, primary care physicians or preoperative screening - received a flyer; 22 households; 81.8% response rate). | ≥18 years old; living with someone with a confirmed positive test of COVID-19. | 55.6%; 43y (range 25-77); na. |
| **Wessely et al., 2022**<sup>53</sup> | Germany; 12 December 2020 to 6 January 2021. | Cross sectional; online survey. | Population based; taken from the CoCo-Fakt cohort study, identified through government register of infected people and their close contacts*, N=8,075 included; 22.7%.<br>*\*data from the same study as Joisten et al. (2022), but reporting different outcomes.* | >16 years of age*<br>Exclusion criteria: noncompliant people; deceased patients; those who were in medical or nursing facilities or quarantined for other reasons (e.g., travel returnees)<br>*\*16 and 17 year olds were included, although the number was not reported, the mean age and SD suggests that they were a small minority and the study was therefore included* | 61.5%; 41.6y (14.2);<br>Infected N=3,208<br>Contacts N=4,867. |
| **Xu et al., 2020**<sup>54</sup> | China; 8 to 21 February 2020. | Cross sectional; online survey. | Population based; survey was 'posted on the Internet' using convenience sampling; N=328; not reported. | ≥18 years old; resident in mainland China. | 39.0%; 31.0y (6.8) range=18-56; na. |

### Aim 1: The impact of self-isolation on psychological wellbeing

Associations between self-isolation and wellbeing were reported in 11 quantitative studies, several of which reported more than one wellbeing outcome. The most reported outcomes were anxiety symptoms,^27,36,39,42,46,50,51^ depressive symptoms,^27,36,39,42,50,51^ general psychological symptoms,^19,20,25,27,46^ post-traumatic stress disorder (PTSD),^23,50^ and stress related symptoms (Table 2).^36,50^ Several other outcomes were reported by only one study, such as loneliness and substance use, which are not synthesised here but can be found in the full extraction tables (Appendix 6). Evidence was inconsistent; grouping by lockdown context did not alter the pattern of results. Risk of bias is summarised and then the outcomes reported in the highest number of studies are discussed first.

**Table 2.** The impact of self-isolation on psychological symptoms and/or diagnosis.

|  | Anxiety | Depressive | General psychological | PTSD | Stress |
| --- | --- | --- | --- | --- | --- |
| Isolating (yes) | ↔<sup>27</sup>↔<sup>39</sup>↑<sup>50</sup>↔<sup>4</sup><br>6↔<sup>36</sup>↔<sup>42</sup>↑<sup>51</sup> | ↑<sup>27</sup>↔<sup>39</sup>↔<sup>50</sup>↔<sup>3</sup><br>6↔<sup>42</sup>↑<sup>51</sup> | ↔<sup>19</sup>↔<sup>27</sup>↔<sup>39</sup>↑<sup>2</sup><br>0↑<sup>46</sup>↑<sup>25</sup> | ↑<sup>23</sup>↑<sup>50</sup> | ↔<sup>50</sup>↔<sup>36</sup> |

Note. Up/down arrow indicates a significant increase/decrease of symptoms in the isolating group, horizontal arrow indicates no identified significant difference, effect arrows are ordered by risk of bias; \*arrow indicates direction of effect in the infected group; \*\*arrow indicates direction of effect in the home group; Risk of bias = ROBINS-E (Exposure) – orange colour = some concerns, red colour = high risk, black colour = very high risk

In the risk of bias assessments, no quantitative findings were low risk. Five findings had some concerns,^19,23,27,39,50^ five were high risk,^20,36,42,46,51^, and one was very high risk.^25^ Only one of the four qualitative studies scored more than 40% on the quality appraisal tool,^52^ and even the higher-quality study was at substantial risk of bias due to the authors’ analysis using an a priori framework based on ‘areas of interest’. The risk of bias summaries are reported in Appendix 7.

For anxiety and depressive symptoms, most studies found no evidence of an effect of self-isolation, (^27,36,39,42,46^ and ^36,39,42,50^ respectively) while two reported worse symptoms in those who had self-isolated (^50,51^ and ^27,51^ respectively). Limiting findings to studies at lowest risk of bias (some concerns),^27,39,50^ did not change the pattern of findings.

For general psychological symptoms, three studies found no evidence for an association with self-isolation.^19,27,39^ Two studies that reported worse general psychological symptoms in those who had self-isolated were carried out under rapidly changing societal contexts.^20,25^ Limiting findings to studies at lowest risk of bias (some concerns),^19,27,39^ suggested no evidence for an effect of self-isolation.

Studies on PTSD symptoms consistently reported a positive association with self-isolation, both were large population cohort studies early in the pandemic, and both had some concerns of bias.^23,50^ Two studies found no evidence for an association between self-isolation and with stress, also early in the pandemic, at high risk and some concerns of bias respectively.^36,50^

Evidence for an impact of self-isolation on psychological symptoms was more consistent in qualitative studies, (Table 3). Participants also described feeling lonely,^26,28,33^ sad,^26,28^ angry and frustrated,^26,52^ bored,^28,33,38,52^ and afraid.^33,52^ They also reported negative impacts on their family relationships.^26,38^ In contrast, some also reported self-isolation to be relaxing and providing more time with family.^28,38^

**Table 3.**
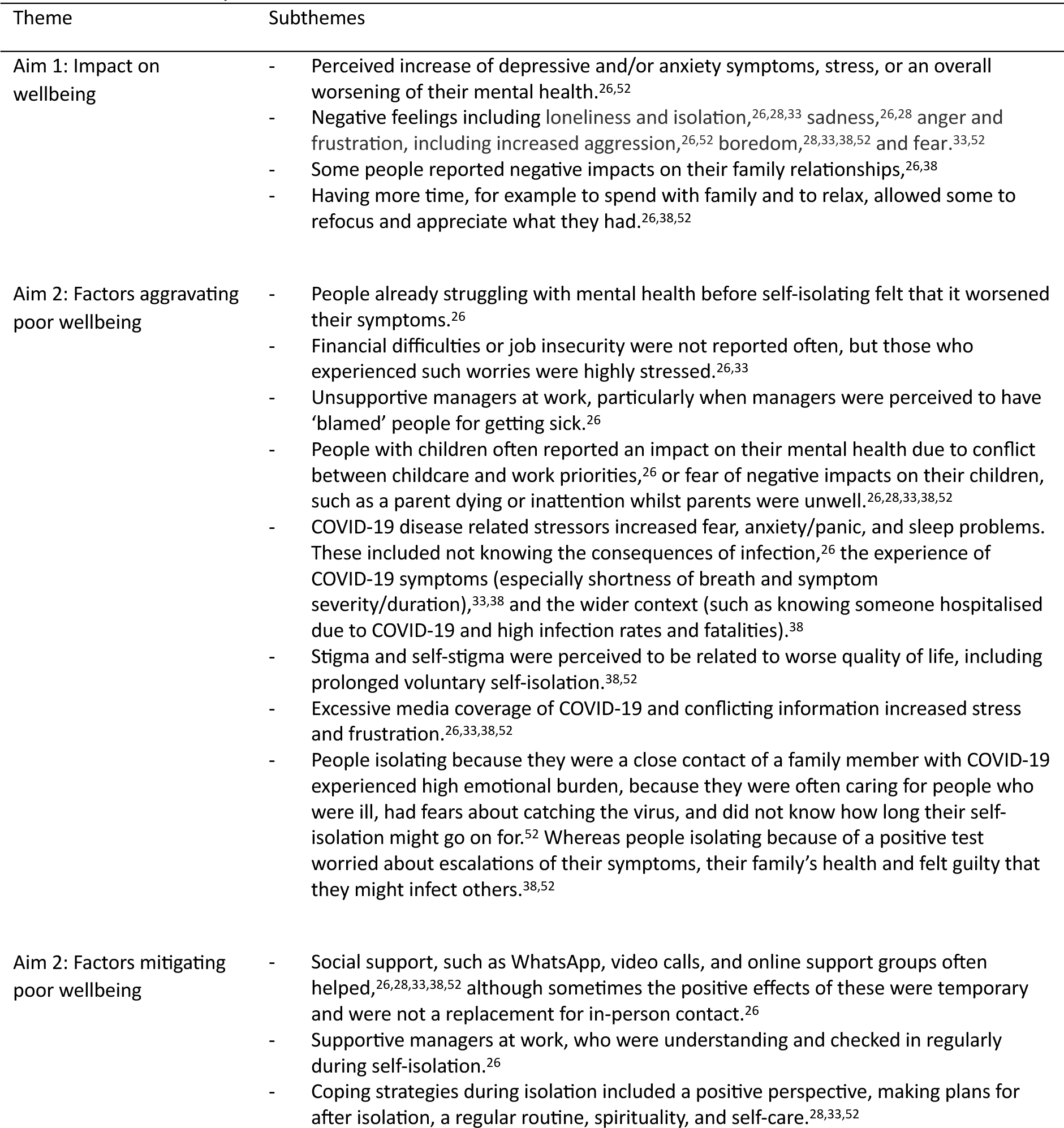
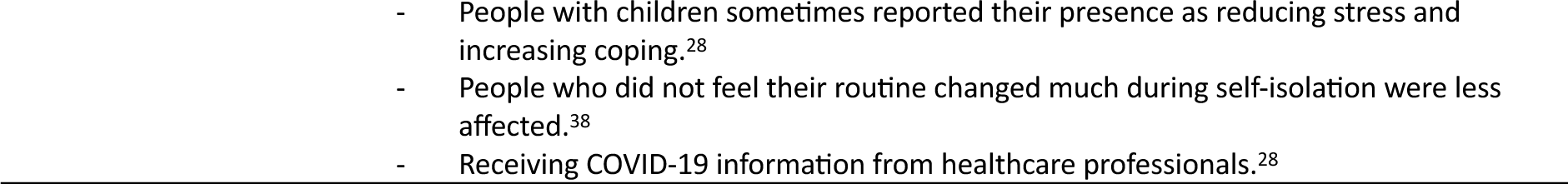
Qualitative synthesised results.

| Theme | Subthemes |
| --- | --- |
| Aim 1: Impact on wellbeing | - Perceived increase of depressive and/or anxiety symptoms, stress, or an overall worsening of their mental health.<sup>26,52</sup><br>- Negative feelings including loneliness and isolation,<sup>26,28,33</sup> sadness,<sup>26,28</sup> anger and frustration, including increased aggression,<sup>26,52</sup> boredom,<sup>28,33,38,52</sup> and fear.<sup>33,52</sup><br>- Some people reported negative impacts on their family relationships,<sup>26,38</sup><br>- Having more time, for example to spend with family and to relax, allowed some to refocus and appreciate what they had.<sup>26,38,52</sup> |
| Aim 2: Factors aggravating poor wellbeing | - People already struggling with mental health before self-isolating felt that it worsened their symptoms.<sup>26</sup><br>- Financial difficulties or job insecurity were not reported often, but those who experienced such worries were highly stressed.<sup>26,33</sup><br>- Unsupportive managers at work, particularly when managers were perceived to have 'blamed' people for getting sick.<sup>26</sup><br>- People with children often reported an impact on their mental health due to conflict between childcare and work priorities,<sup>26</sup> or fear of negative impacts on their children, such as a parent dying or inattention whilst parents were unwell.<sup>26,28,33,38,52</sup><br>- COVID-19 disease related stressors increased fear, anxiety/panic, and sleep problems. These included not knowing the consequences of infection,<sup>26</sup> the experience of COVID-19 symptoms (especially shortness of breath and symptom severity/duration),<sup>33,38</sup> and the wider context (such as knowing someone hospitalised due to COVID-19 and high infection rates and fatalities).<sup>38</sup><br>- Stigma and self-stigma were perceived to be related to worse quality of life, including prolonged voluntary self-isolation.<sup>38,52</sup><br>- Excessive media coverage of COVID-19 and conflicting information increased stress and frustration.<sup>26,33,38,52</sup><br>- People isolating because they were a close contact of a family member with COVID-19 experienced high emotional burden, because they were often caring for people who were ill, had fears about catching the virus, and did not know how long their self-isolation might go on for.<sup>52</sup> Whereas people isolating because of a positive test worried about escalations of their symptoms, their family's health and felt guilty that they might infect others.<sup>38,52</sup> |
| Aim 2: Factors mitigating poor wellbeing | - Social support, such as WhatsApp, video calls, and online support groups often helped,<sup>26,28,33,38,52</sup> although sometimes the positive effects of these were temporary and were not a replacement for in-person contact.<sup>26</sup><br>- Supportive managers at work, who were understanding and checked in regularly during self-isolation.<sup>26</sup><br>- Coping strategies during isolation included a positive perspective, making plans for after isolation, a regular routine, spirituality, and self-care.<sup>28,33,52</sup> |

### Aim 2: Factors associated with psychological wellbeing during or following self-isolation

Factors associated with wellbeing were reported in 20 quantitative studies, several of which investigated associations with more than one factor (Table 4). The most reported factors were related to self-isolation,^19,21,32,34,35,37,40,44,45,49,51^ demographics,^21,30,32,42,44,47^ mental and physical health,^21,32,37,47,54^ and COVID-19 symptoms.^21,37,43^ Several factors or outcomes were reported by only one study, such as the time of year or loneliness, which are not reported here but can be found in the full extraction tables (Appendix 6). Evidence was often inconsistent. A general narrative summary is provided here; full details are in Appendix 8.

**Table 4.** Factors associated with psychological symptoms and/or diagnosis during self-isolation.

| Predictor type | Predictor | Anxiety | Depressive | General psychological | PTSD | Insomnia |
| --- | --- | --- | --- | --- | --- | --- |
| Isolation | Infected vs close contact* | ↔<sup>45</sup>↔<sup>19</sup> | ↑<sup>45</sup>↑<sup>19</sup> | ↔<sup>19</sup>↑<sup>34</sup> |  |  |
|  | Home vs hotel/hospital** | ↔<sup>45</sup>↔<sup>51</sup>↔<sup>40</sup> | ↓<sup>45</sup>↔<sup>49</sup>↔<sup>51</sup>↑<sup>40</sup> |  | ↓<sup>49</sup>↓<sup>40</sup> |  |
|  | Duration (end versus start of self-isolation) | ↓<sup>35</sup> | ↓<sup>35</sup> |  |  | ↓<sup>21</sup> |
|  | Covid-related stressors (yes) | ↑<sup>44</sup> | ↑<sup>32</sup>↑<sup>44</sup> | ↑<sup>37</sup> |  |  |
|  | Assistance/support (no) |  | ↑<sup>32</sup> | ↑<sup>37</sup> |  |  |
| Demographic | Age (younger) |  | ↑<sup>32</sup> | ↓<sup>30</sup> |  | ↔<sup>21</sup> |
|  | Gender (female) | ↔<sup>42</sup>↑<sup>44</sup>↑<sup>47</sup> | ↔<sup>32</sup>↔<sup>42</sup>↑<sup>44</sup> | ↑<sup>30</sup> |  |  |
|  | Living alone (yes) |  | ↔<sup>32</sup> | ↔<sup>30</sup> |  |  |
| Covid-symptoms | Viral load / covid symptoms (higher) | ↔<sup>43</sup> | ↔<sup>43</sup> | ↑<sup>37</sup>↑<sup>43</sup> |  | ↔<sup>21</sup>↔<sup>43</sup> |
| Mental/physical health | Psychological symptoms (any; yes/higher) | ↑<sup>54</sup>↑<sup>47</sup> |  | ↑<sup>37</sup> |  | ↑<sup>21</sup> |
|  | Poor physical health (yes) |  | ↑<sup>32</sup> | ↑<sup>37</sup> |  |  |
|  | Coping strategies (no) | ↑<sup>54</sup> |  | ↑<sup>37</sup> |  |  |

Note. Up/down arrow indicates a significant increase/decrease of symptoms in the isolating group, horizontal arrow indicates no identified significant difference, effect arrows are ordered by risk of bias; \*arrow indicates direction of effect in the infected group; \*\*arrow indicates direction of effect in the home group; Risk of bias = ROBINS-E (Exposure) – orange colour = some concerns, red colour = high risk, black colour = very high risk.

In risk of bias assessments, no findings were at low risk, one had some concerns,^35^ and most findings were at high,^21,30,32,41,42,44,45,49,51,53^ or very high risk of bias.^19,22,29,34,37,40,43,47,54^; (Appendix 7). Limiting findings to some concerns of bias or high risk of bias did not alter the interpretation of the findings for aim 2.

Factors related to self-isolation were reported in eleven studies. COVID-19 stressors, such as changes in daily life, worries about infection and job security, and a lack of support during self-isolation were consistently associated with higher levels of general psychological, depressive, and anxiety symptoms.^32,37,44^ Depressive and anxiety symptoms were found to be lower at the end of self-isolation compared to the start ^35^ and a longer period of self-isolation was associated with lower sleep problems at the end of the isolation period.^21^ Self-isolation at home rather than a hotel was associated with lower PTSD symptoms,^40,49^ but not anxiety symptoms.^40,45,51^

Factors related to demographics were reported in six studies. There was no evidence found for living alone to be associated with wellbeing symptoms.^30,32^ There was inconsistent evidence for an association between age or gender with any wellbeing symptoms.^21,30,32,42,44,47^

Factors related to mental or physical health were reported in five studies, all of which reported associations with wellbeing outcomes including psychological burden,^37^ depressive and anxiety symptoms,^32,47,54^ and insomnia.^21^ Low levels of coping strategies were associated with psychological burden,^37^ and anxiety symptoms.^54^

Factors related to COVID-19 were reported in three studies, including higher viral load and more severe symptoms.^21,37,43^ Both these factors were associated with an increase in general psychological symptoms.^37,43^ Whereas no evidence was found for an association with depressive or anxiety symptoms, or insomnia.^21,43^

Qualitative findings largely supported the associations identified in the quantitative findings (Table 3). Other factors which participants perceived to increase the negative wellbeing impacts of self-isolation included fears around COVID-19,^26,33,38,52^ financial difficulties,^26,33^ stigma,^38,52^ exposure to excessive media coverage of COVID-19,^33^ and conflicting guidance on how to isolate and symptom prognosis.^26,38,52^ Factors which participants perceived to reduce the negative impact of self-isolation on wellbeing included social support,^26,28,33,38,52^ coping strategies such as making plans for after isolation, keeping a regular routine, spirituality, and self-care,^28,33,52,38^ and receiving reliable information about COVID-19 from healthcare professionals.^28^ In addition, some people also reported a positive impact on their wellbeing, such as having more time to spend with family and to relax, which allowed them to refocus and appreciate what they had.^26,38,52^

Qualitative findings also indicated that the same factor could have a positive or negative impact on wellbeing, depending on the person’s individual context. For example, having children,^26,28^ or uncertainties around symptoms and duration.^26,33,38^

### Aim 3: Interventions to improve psychological wellbeing during self-isolation

Three studies investigated the effect of an intervention on wellbeing. ^24,31,48^ One study tested a telenursing intervention, reporting a greater reduction in anxiety symptoms from pre-test to post-test in the intervention group compared to the control group. ^24^ Two studies tested yogic meditation interventions, reporting a decrease in depressive and general psychological symptoms, and insomnia, and an increase in adaptive wellbeing, at the end of the intervention period compared to the start (Table 5). ^31,48^ Both meditation intervention studies were conducted in the same hospital, using the same outcomes and similar interventions, but at least partially different participants. Study quality was problematic for all interventions, which rated as high risk, ^24^ or very high risk (Appendix 7). ^31,48^

**Table 5.** The impact of interventions to improve psychological symptoms and/or diagnosis during self-isolation.

| Intervention | Anxiety | Depressive | Stress | Insomnia | Adaptive wellbeing |
| --- | --- | --- | --- | --- | --- |
| Telenursing | ↓<sup>24</sup> |  |  |  |  |
| Yogic meditation* | ↓<sup>31</sup>↔<sup>48</sup> | ↓<sup>31</sup>↓<sup>48</sup> | ↓<sup>31</sup>↔<sup>48</sup> | ↓<sup>31</sup>↓<sup>48</sup> | ↑<sup>31</sup>↑<sup>48</sup> |

Note. Up/down arrow indicates a significant increase/decrease of symptoms in the isolating group, horizontal arrow indicates no identified significant difference, effect arrows are ordered by risk of bias; \*arrow indicates direction of effect in the infected group; \*\*arrow indicates direction of effect in the home group; Risk of bias = ROBINS-I (Intervention) – red colour = high risk, black colour = very high risk.

## Discussion

This systematic review summarises the literature on self-isolation and psychological wellbeing during the COVID-19 pandemic. Overall, there was considerable heterogeneity in wellbeing outcomes and isolation contexts reported in the studies. Most studies were cross-sectional, survey based, and at high or very high risk of bias. Quantitative evidence for an association between self-isolation and wellbeing was inconsistent, although some clear associations emerged for specific outcomes such as increased PTSD symptoms. Qualitative evidence showed a more consistent negative impact of self-isolation. Stressors, including pre-existing health needs and low levels of support, were consistently associated with worse wellbeing outcomes. Intervention studies were rare and at high or very high risk of bias.

Quantitative studies largely reported psychopathology symptom scales, which may explain the discrepancy between their findings and those of qualitative studies. These scales rarely assessed broader aspects of wellbeing such as worry, stigma, and somatic pain that could lead to behavioural changes like obsessive protective behaviours lasting far beyond the period of self-isolation, as found in other infectious disease contexts. ^26,38,55,56^ These broader aspects of wellbeing, and others such as frustration and agitation, may also have broader consequences, including lowering trust in governments and reducing adherence to mitigation measures. ^57,58^ Future research should consider wellbeing beyond psychopathology symptoms to better characterise the impact of self-isolation on wellbeing, whilst also continuing to focus on identifying groups with symptom levels suggestive of a treatment need.

One exception was PTSD symptoms, which were consistently higher in people who were currently or previously in self-isolation. ^23,50^ This is consistent with a previous review that found increased levels of PTSD symptoms in quarantined individuals across all contexts, including different infectious diseases and different groups such as parents, the general population, and healthcare workers. ^2^ Additionally, home isolation was consistently associated with reduced PTSD symptoms compared to isolation in a managed facility. ^40,49^ These results suggest that home isolation should be prioritised to reduce the risk of PTSD symptoms during self-isolation.

Self-isolation was found to associate with wellbeing differently, depending on individual factors and context. There was good evidence that people with greater mental and physical health needs, ^26,29,32,37,44,47,54^ who experienced COVID-related stressors including inadequate support, ^26,32,33,37,38,44,52^ and had reduced coping strategies, ^37,53,54^ were most at risk of adverse outcomes. As found during COVID-19 lockdowns, ^59,60^ parents and carers were more at risk, especially when there was conflict between childcare and work. ^26^ However, the presence of children could also reduce stress and helped some parents to cope. ^28^ This complexity mirrors the intricacies observed in prior studies, which found that factors such as the number and age of the children could exert different risk or protective effects. ^61,62^ Future research should focus on identifying the subpopulations most at risk of adverse wellbeing outcomes during self-isolation and developing and evaluating targeted public health interventions to support these groups, including providing practical support and promoting coping strategies to those who need it.

Few studies investigating the effect of interventions on wellbeing during self-isolation were identified. All three studies reported a supposed impact of the intervention on most wellbeing outcomes, ^24,31,48^ but several non-intervention studies also found that people generally experienced a reduction of symptoms over the course of isolation^21,35^ This suggests that two of the intervention studies, which lacked control groups, may have overestimated the effect of the intervention. Nevertheless, the study that used a control group found a greater reduction in anxiety at the end of the intervention (i.e. at follow-up) in those who received daily tele-nursing. ^24^ Qualitative studies highlighted modifiable stressors that have been consistently reported in previous infectious disease and disaster contexts, such as inadequate and conflicting information, ^55,63,64^ leading to heightened fears of disease progression and extended isolation. ^65,66^ Together, these findings suggest that high-quality intervention studies should be prioritised to better understand how to mitigate the impact of self-isolation on wellbeing. Emphasis should be placed on interventions targeting loneliness and misinformation, for example, through regular contact with healthcare professionals, whilst monitoring and identifying individuals who may require additional support.

### Strengths and limitations

To our knowledge, this is the first review to explore the relationship between self-isolation and psychological wellbeing during the COVID-19 pandemic. Strengths include a pre-registered protocol, no geographical or language limitations, comprehensive risk of bias assessments, a robust process for agreement, and adherence to PRISMA, SWiM, and eMERGe guidelines. Limitations of the studies were the inconsistent use of the terms ‘isolation’ and ‘quarantine’, which could have led to the exclusion of relevant research, and the high risk of bias in many studies. Limitations of the review were that we were unable to formally analyse publication bias and did not examine heterogeneity in study design in the synthesis, which limits our confidence in the certainty of the evidence presented. Human error may have resulted in missed studies.

### Implications and conclusions

Self-isolation impacts psychological wellbeing, particularly for PTSD symptoms. Self-isolating at home may reduce this risk, but more and better-quality evidence is needed. A significant limitation of the research base is its over reliance on psychopathology symptom questionnaires, which may miss the substantial impact of self-isolation on broader aspects of wellbeing, such as frustration or stigma. Ignoring these impacts can mean that potential downstream effects such as trust in government and reduced adherence to mitigation measures are overlooked. Public health officials should make it a priority to support vulnerable individuals with pre-existing health conditions, lack of support, or significant life stressors when implementing self-isolation directives in the future. Clinicians and healthcare workers can play a key role in identifying and supporting those most at risk. Interventions should focus on addressing loneliness, worries, and misinformation, and monitoring and identifying individuals who need additional support.

## Contributors

AFM, LES, SKB, and GJR conceptualised the study. AFM ran the systematic searches and curated the data. AFM, LES, SKB, RD, MVS and GJR screened citations. AFM and SKB completed data extraction and risk of bias ratings. AFM wrote the original draft. LES, SKB, RD, MVS, RA, NG, and GJR reviewed, edited, and approved the final manuscript. All authors were involved in the scientific processes leading up to the writing of the manuscript and contributed to reviewing the papers, the interpretation of the findings, and the critical evaluation of the final version of the manuscript. All authors had full access to the data and accept responsibility to submit for publication. LES and AFM are guarantors. The corresponding author attests that all listed authors meet authorship criteria and that no others meeting the criteria have been omitted.

## Data sharing

No novel data were collected as part of this study. All data are already publicly available.

## Data Availability

No novel data were collected as part of this study. All data are already publicly available.

## Acknowledgements

This study was funded by the Research England *Policy Support Fund* 2022-23 (from the allocation to King’s College London). AFM, LES, SKB, RA and GJR are supported by the National Institute for Health and Care Research Health Protection Research Unit (NIHR HPRU) in Emergency Preparedness and Response, a partnership between the UK Health Security Agency, King’s College London and the University of East Anglia. The views expressed are those of the authors and not necessarily those of the NIHR, UKHSA or the Department of Health and Social Care. For the purpose of open access, the author has applied a Creative Commons Attribution (CC BY) licence to any Author Accepted Manuscript version arising.

## AI statement

During the preparation of this work, AI-assisted technologies were used in the writing process to improve readability and language of the work. AI was not used to produce scientific insights, analyse or interpret data, or draw scientific conclusions. Application of AI was performed with human oversight and control. The authors reviewed and edited the content, and are responsible and accountable for the originality, accuracy, and integrity of the work.

## Ethics

All data used were in the public domain, therefore ethical approval was not required.

