## Supplement for "The impact of self-isolation on psychological wellbeing and how to reduce it: a systematic review"

**Appendix 1: Reporting Checklists**

1. *Prisma 2020 for Abstracts Checklist*

| Section and Topic | Item # | Checklist item | Reported y/n |
|---|---|---|---|
| **TITLE** | | | |
| Title | 1 | Identify the report as a systematic review. | Yes |
| **BACKGROUND** | | | |
| Objectives | 2 | Provide an explicit statement of the main objective(s) or question(s) the review addresses. | Yes |
| **METHODS** | | | |
| Eligibility criteria | 3 | Specify the inclusion and exclusion criteria for the review. | Yes |
| Information sources | 4 | Specify the information sources (e.g. databases, registers) used to identify studies and the date when each was last searched. | Yes |
| Risk of bias | 5 | Specify the methods used to assess risk of bias in the included studies. | Yes |
| Synthesis of results | 6 | Specify the methods used to present and synthesise results. | Yes |
| **RESULTS** | | | |
| Included studies | 7 | Give the total number of included studies and participants and summarise relevant characteristics of studies. | Yes |
| Synthesis of results | 8 | Present results for main outcomes, preferably indicating the number of included studies and participants for each. If meta-analysis was done, report the summary estimate and confidence/credible interval. If comparing groups, indicate the direction of the effect (i.e. which group is favoured). | Yes |
| **DISCUSSION** | | | |
| Limitations of evidence | 9 | Provide a brief summary of the limitations of the evidence included in the review (e.g. study risk of bias, inconsistency and imprecision). | Yes |
| Interpretation | 10 | Provide a general interpretation of the results and important implications. | Yes |
| **OTHER** | | | |
| Funding | 11 | Specify the primary source of funding for the review. | Yes |
| Registration | 12 | Provide the register name and registration number. | Yes |

From: Page, M. J., McKenzie, J. E., Bossuyt, P. M., Boutron, I., Hoffmann, T. C., Mulrow, C. D., Shamseer, L., Tetzlaff, J. M., Akl, E. A., Brennan, S. E., Chou, R., Glanville, J., Grimshaw, J. M., Hrobjartsson, A., Lalu, M. M., Li, T., Loder, E. W., Mayo-Wilson, E., McDonald, S., . . . Moher, D. (2021). The PRISMA 2020 statement: An updated guideline for reporting systematic reviews. *Int J Surg*, *88*, 105906. https://doi.org/10.1016/j.ijsu.2021.105906

*2. PRISMA 2020 (Study) checklist*

| **Section and Topic** | **Item #** | **Checklist item** | **Location where item is reported** |
|---|---|---|---|
| **TITLE** | | | |
| Title | 1 | Identify the report as a systematic review. | Page 1 |
| **ABSTRACT** | | | |
| Abstract | 2 | See the PRISMA 2020 for Abstracts checklist. | Appendix Page 2 |
| **INTRODUCTION** | | | |
| Rationale | 3 | Describe the rationale for the review in the context of existing knowledge. | Page 3 |
| Objectives | 4 | Provide an explicit statement of the objective(s) or question(s) the review addresses. | Page 4 |
| **METHODS** | | | |
| Eligibility criteria | 5 | Specify the inclusion and exclusion criteria for the review and how studies were grouped for the syntheses. | Pages 4-5; Appendix 3 |
| Information sources | 6 | Specify all databases, registers, websites, organisations, reference lists and other sources searched or consulted to identify studies. Specify the date when each source was last searched or consulted. | Page 4; Appendix 2 |
| Search strategy | 7 | Present the full search strategies for all databases, registers and websites, including any filters and limits used. | Page 5; Appendix 2 |
| Selection process | 8 | Specify the methods used to decide whether a study met the inclusion criteria of the review, including how many reviewers screened each record and each report retrieved, whether they worked independently, and if applicable, details of automation tools used in the process. | Pages 4-5; Appendix 3 |
| Data collection | 9 | Specify the methods used to collect data from reports, including how many reviewers collected data from each report, whether they worked independently, any processes for obtaining or confirming data | Page 5 |

| Section and Topic | Item # | Checklist item | Location where item is reported |
|---|---|---|---|
| process | | from study investigators, and if applicable, details of automation tools used in the process. | |
| Data items | 10a | List and define all outcomes for which data were sought. Specify whether all results that were compatible with each outcome domain in each study were sought (e.g. for all measures, time points, analyses), and if not, the methods used to decide which results to collect. | Page 5, Tables 2,5 & 6; Appendix 6, Table 3 |
| | 10b | List and define all other variables for which data were sought (e.g. participant and intervention characteristics, funding sources). Describe any assumptions made about any missing or unclear information. | Page 5; Table 1; Appendix 6, Tables 1 & 2 |
| Study risk of bias assessment | 11 | Specify the methods used to assess risk of bias in the included studies, including details of the tool(s) used, how many reviewers assessed each study and whether they worked independently, and if applicable, details of automation tools used in the process. | Pages 5-6; Appendix 4 |
| Effect measures | 12 | Specify for each outcome the effect measure(s) (e.g. risk ratio, mean difference) used in the synthesis or presentation of results. | Tables 2,4 & 5 |
| Synthesis methods | 13a | Describe the processes used to decide which studies were eligible for each synthesis (e.g. tabulating the study intervention characteristics and comparing against the planned groups for each synthesis (item #5)). | Page 6; Appendix 5 |
| | 13b | Describe any methods required to prepare the data for presentation or synthesis, such as handling of missing summary statistics, or data conversions. | Page 6; Appendix 5 |
| | 13c | Describe any methods used to tabulate or visually display results of individual studies and syntheses. | Page 6; Appendix 5 |
| | 13d | Describe any methods used to synthesize results and provide a rationale for the choice(s). If meta-analysis was performed, describe the model(s), method(s) to identify the presence and extent of statistical heterogeneity, and software package(s) used. | Page 6; Appendix 5 |
| | 13e | Describe any methods used to explore possible causes of heterogeneity among study results (e.g. subgroup analysis, meta-regression). | Page 17 & 21; Appendix 5 |
| | 13f | Describe any sensitivity analyses conducted to assess robustness of the synthesized results. | n/a |

| Section and Topic | Item # | Checklist item | Location where item is reported |
|---|---|---|---|
| Reporting bias assessment | 14 | Describe any methods used to assess risk of bias due to missing results in a synthesis (arising from reporting biases). | Appendix 7 |
| Certainty assessment | 15 | Describe any methods used to assess certainty (or confidence) in the body of evidence for an outcome. | Pages 18-23 |
| **RESULTS** | | | |
| Study selection | 16a | Describe the results of the search and selection process, from the number of records identified in the search to the number of studies included in the review, ideally using a flow diagram. | Pages 7-8 |
| | 16b | Cite studies that might appear to meet the inclusion criteria, but which were excluded, and explain why they were excluded. | Pages 7-8; Appendix 3 |
| Study characteristics | 17 | Cite each included study and present its characteristics. | Pages 10-17 |
| Risk of bias in studies | 18 | Present assessments of risk of bias for each included study. | Appendix 7 |
| Results of individual studies | 19 | For all outcomes, present, for each study: (a) summary statistics for each group (where appropriate) and (b) an effect estimate and its precision (e.g. confidence/credible interval), ideally using structured tables or plots. | Tables 2-5 |
| Results of syntheses | 20a | For each synthesis, briefly summarise the characteristics and risk of bias among contributing studies. | Pages 17-22; Appendix 7 |
| | 20b | Present results of all statistical syntheses conducted. If meta-analysis was done, present for each the summary estimate and its precision (e.g. confidence/credible interval) and measures of statistical heterogeneity. If comparing groups, describe the direction of the effect. | Pages 17-22; Appendix 8 |
| | 20c | Present results of all investigations of possible causes of heterogeneity among study results. | Pages 17-22; Appendices 5 & 8 |
| | 20d | Present results of all sensitivity analyses conducted to assess the robustness of the synthesized results. | n/a |
| Reporting biases | 21 | Present assessments of risk of bias due to missing results (arising from reporting biases) for each | Appendix 7 |

| Section and Topic | Item # | Checklist item | Location where item is reported |
|---|---|---|---|
| | | synthesis assessed. | |
| Certainty of evidence | 22 | Present assessments of certainty (or confidence) in the body of evidence for each outcome assessed. | Pages 17-22 |
| **DISCUSSION** | | | |
| Discussion | 23a | Provide a general interpretation of the results in the context of other evidence. | Pages 22-24 |
| | 23b | Discuss any limitations of the evidence included in the review. | Page 24 |
| | 23c | Discuss any limitations of the review processes used. | Page 24 |
| | 23d | Discuss implications of the results for practice, policy, and future research. | Page 24 |
| **OTHER INFORMATION** | | | |
| Registration and protocol | 24a | Provide registration information for the review, including register name and registration number, or state that the review was not registered. | Pages 1 & 4 |
| | 24b | Indicate where the review protocol can be accessed, or state that a protocol was not prepared. | Pages 1 & 4 |
| | 24c | Describe and explain any amendments to information provided at registration or in the protocol. | Appendix 3 |
| Support | 25 | Describe sources of financial or non-financial support for the review, and the role of the funders or sponsors in the review. | Page 3 & 25 |
| Competing interests | 26 | Declare any competing interests of review authors. | Page 25 |
| Availability of data, code and other materials | 27 | Report which of the following are publicly available and where they can be found: template data collection forms; data extracted from included studies; data used for all analyses; analytic code; any other materials used in the review. | |

From: Page, M. J., McKenzie, J. E., Bossuyt, P. M., Boutron, I., Hoffmann, T. C., Mulrow, C. D., Shamseer, L., Tetzlaff, J. M., Akl, E. A., Brennan, S. E., Chou, R., Glanville, J., Grimshaw, J. M., Hrobjartsson, A., Lalu, M. M., Li, T., Loder, E. W., Mayo-Wilson, E., McDonald, S., . . . Moher, D. (2021). The PRISMA 2020 statement: An updated guideline for reporting systematic reviews. *Int J Surg*, *88*, 105906. https://doi.org/10.1016/j.ijsu.2021.105906

*3. Synthesis Without Meta-analysis (SWiM) reporting items*

| **SWiM is intended to complement and be used as an extension to PRISMA** | | | |
|---|---|---|---|
| **SWiM reporting item** | **Item description** | **Page in manuscript where item is reported** | **Other*** |
| *Methods* | | | |
| **1** Grouping studies for synthesis | 1a) Provide a description of, and rationale for, the groups used in the synthesis (e.g., groupings of populations, interventions, outcomes, study design) | Page 6; Appendix 5 | |
| | 1b) Detail and provide rationale for any changes made subsequent to the protocol in the groups used in the synthesis | Appendix 3 | |
| **2** Describe the standardised metric and transformation methods used | Describe the standardised metric for each outcome. Explain why the metric(s) was chosen, and describe any methods used to transform the intervention effects, as reported in the study, to the standardised metric, citing any methodological guidance consulted | Page 6; Appendix 5 | |
| **3** Describe the synthesis methods | Describe and justify the methods used to synthesise the effects for each outcome when it was not possible to undertake a meta-analysis of effect estimates | Page 6; Appendix 5 | |
| **4** Criteria used to prioritise results for summary and synthesis | Where applicable, provide the criteria used, with supporting justification, to select the particular studies, or a particular study, for the main synthesis or to draw conclusions from the synthesis (e.g., based on study design, risk of bias assessments, directness in relation to the review question) | Pages 17 & 19 | |

| | | |
|---|---|---|
| **5** Investigation of heterogeneity in reported effects | State the method(s) used to examine heterogeneity in reported effects when it was not possible to undertake a meta-analysis of effect estimates and its extensions to investigate heterogeneity | Pages 17-22; Appendix 5 |
| **6** Certainty of evidence | Describe the methods used to assess certainty of the synthesis findings | Pages 17-22; Appendix 4 |
| **7** Data presentation methods | Describe the graphical and tabular methods used to present the effects (e.g., tables, forest plots, harvest plots). Specify key study characteristics (e.g., study design, risk of bias) used to order the studies, in the text and any tables or graphs, clearly referencing the studies included | Tables 2-5; Appendix 5 |

*Results*

| | | |
|---|---|---|
| **8** Reporting results | For each comparison and outcome, provide a description of the synthesised findings, and the certainty of the findings. Describe the result in language that is consistent with the question the synthesis addresses, and indicate which studies contribute to the synthesis | Pages 17-22; Appendix 6 |

*Discussion*

| | | |
|---|---|---|
| **9** Limitations of the synthesis | Report the limitations of the synthesis methods used and/or the groupings used in the synthesis, and how these affect the conclusions that can be drawn in relation to the original review question | Page 25 |

PRISMA=Preferred Reporting Items for Systematic Reviews and Meta-Analyses.

*If the information is not provided in the systematic review, give details of where this information is available (e.g., protocol, other published papers (provide citation details), or website (provide the URL)).

Campbell, M., McKenzie, J. E., Sowden, A., Katikireddi, S. V., Brennan, S. E., Ellis, S., Hartmann-Boyce, J., Ryan, R., Shepperd, S., Thomas, J., Welch, V., & Thomson, H. (2020). Synthesis without meta-analysis (SWiM) in systematic reviews: reporting guideline. *BMJ*, *368*, l6890. https://doi.org/10.1136/bmj.l6890

**Appendix 2: Search strategy and results**

A systematic search was conducted for studies published between 01 January 2020 and 13 December 2022. The peer-review and preprint searches were carried out between 14-17 December 2022, backward reference checking continued until 16 February 2023, and grey literature searches were conducted until 24 July 2023. We searched six peer-review and preprint databases (Medline, PsycInfo, Web of Science, Embase, PsyArXiv, medRxiv) and five grey literature databases (OpenGrey, World Health Organization [WHO], National Technical Information Service [NTIS; United States Department of Commerce], WorldCat, and Agency for Healthcare Research and Quality [AHRQ]), recommended by the National Institutes of Health Library.[1] A Google search and searches of the websites for relevant agencies and organisations (e.g. the UK Testing Initiatives Evaluation Board, the UK Office for National Statistics) were used to identify other potentially relevant sources.

The search strategy included terms for COVID-19, isolation and quarantine (combined with NOT social isolation), and psychological wellbeing. Broad and specific search terms were used to maximise the detection of eligible studies. The search was also used as the basis of a separate systematic review exploring adherence to self-isolation (link to #pre-registration/protocol), screening for both reviews up to full text stage was performed in parallel.

[1] Literature Search: Databases and Gray Literature. National Institutes of Health Library.

*Database search example*

| # | Query |
|---|---|
| 1 | (coronavirus or covid* or sars-cov-2 or ncov2019).mp. or exp Coronavirus/ or exp COVID-19/ or exp SARS-CoV-2/ |
| 2 | (isolat* or quarantin* or confinement).mp. or exp Patient Isolation/ or exp Quarantine/ |
| 3 | 2 not "social isolation".mp. [mp=title, book title, abstract, original title, name of substance word, subject heading word, floating sub-heading word, keyword heading word, organism supplementary concept word, protocol supplementary concept word, rare disease supplementary concept word, unique identifier, synonyms] |
| 4 | (adheren* or compliance or wellbeing or well-being or "quality of life" or resilien* or coping or flourish* or "positive psychology" or "posttraumatic growth" or "post-traumatic growth" or "life satisfaction" or "personal satisfaction" or "psychosocial functioning" or "mental health" or anxiety or depress* or ptsd or trauma* or psychiatric or "psychological stress" or "social stigma" or distress* or mood* or emotion* or "substance abuse" or "substance misuse" or "substance use" or "hazardous drinking" or "alcohol use" or "alcohol abuse" or "alcohol misuse" or alcoholi* or sleep or insomnia or loneliness).mp. or exp Guideline Adherence/ or exp "Treatment Adherence and Compliance"/ or exp Compliance/ or exp Patient Compliance/ or exp "Quality of Life"/ or exp Resilience, Psychological/ or exp Psychology, Positive/ or exp Posttraumatic Growth, Psychological/ or exp Personal Satisfaction/ or exp Psychosocial Functioning/ or exp Mental Health/ or exp Anxiety Disorders/ or exp Anxiety/ or exp Panic/ or exp Panic Disorder/ or exp Depression/ or exp Stress Disorders, Post-Traumatic/ or exp Psychological Trauma/ or exp Stress, Psychological/ or exp Social Stigma/ or Psychological Distress/ or exp Emotions/ or exp Sleep/ or exp "Sleep Initiation and Maintenance Disorders"/ or exp Substance Abuse, Intravenous/ or exp Substance-Related Disorders/ or exp Alcoholism/ or exp Alcohol Drinking/ or exp Loneliness/ |
| 5 | 1 and 3 and 4 |
| 6 | limit 5 to (yr="2020 -Current") |

The search functions on the pre-print registers are not suitable for use in systematic reviews due to several limitations (specifically, confusing Boolean operators, a lack of reproducibility, and no batch export). A discussion and code are provided in this blog: https://ropensci.org/blog/2020/10/20/searching-medrxivr-and-biorxiv-preprint-data

To overcome this, preprint searches were extracted using the R package medrxivr. McGuinness, L. A., & Schmidt, L. (2020). medrxivr: Accessing and searching medRxiv and bioRxiv preprint data in R. *Journal of Open Source Software*, *5*(54), 2651.

The code is available on request from the corresponding author.

*Grey literature searches*

Website search (reported on the right-hand side of the PRISMA flow diagram in the main text).

| Number of websites | Website name | Website link(s) | Reports sought for retrieval | Reports not retrieved | Reports assessed for eligibility | Reports included | Reports excluded |
|---|---|---|---|---|---|---|---|
| **1** | Rijksinstituut voor Volksgezondheid en Milieu (Dutch National Institute for Public Health and the Environment) | https://www.rivm.nl/en/coronavirus-covid-19/research/behaviour | 21 | 0 | 21 | 0 | 21 |
| **2** | iCARE (international COVID-19 Awareness and Responses Evaluation) Study | https://icare.mbmc-cmcm.ca/results-findings/results-1/ | 1 | 0 | 1 | 0 | 1 |
| **3** | | https://www.mbmc-cmcm.ca/2021/covid19/results-findings/infographics/ | 1 | 0 | 1 | 0 | 1 |
| **4** | | https://www.mbmc-cmcm.ca/2021/covid19/results-findings/publications/ | 0 | 0 | 0 | 0 | 0 |
| **5** | CHARIS (Covid Health and Adherence Research In Scotland) Study | https://www.abdn.ac.uk/iahs/research/health-psychology/publications-documents-2174.php | 26 | 0 | 26 | 0 | 26 |
| | | **TOTAL:** | **49** | **0** | **49** | **0** | **49** |

Organisation search (reported on the right-hand side of the PRISMA flow diagram in the main text).

| N of organisations | Organisation | Website link(s) | Reports sought for retrieval | Reports not retrieved | Reports assessed for eligibility | Reports included | Reports excluded |
|---|---|---|---|---|---|---|---|
| **1** | Public Health Wales | https://phw.nhs.wales/ | 7 | 0 | 7 | 0 | 7 |
| **2** | HSC Public Health Agency (Northern Ireland) | https://www.publichealth.hscni.net/ | 0 | 0 | 0 | 0 | 0 |
| **3** | NI Direct [GOV] | https://www.nidirect.gov.uk/ | 0 | 0 | 0 | 0 | 0 |
| **4** | Office for National Statistics | https://www.ons.gov.uk/ | 0 | 0 | 0 | 0 | 0 |
| **5** | Welsh Government | https://www.gov.wales/ | 4 | 0 | 4 | 0 | 4 |
| **6** | StatsWales | https://statswales.gov.wales/Catalogue | 0 | 0 | 0 | 0 | 0 |
| **7** | Scottish Government | https://www.gov.scot/ | 16 | 0 | 16 | 0 | 16 |
| **8** | Northern Ireland Executive | https://www.northernireland.gov.uk/publications | 0 | 0 | 0 | 0 | 0 |
| **9** | Department of Health, Northern Ireland | https://www.health-ni.gov.uk/covid-19-statistics | 0 | 0 | 0 | 0 | 0 |
| **10** | Northern Ireland Statistics and Research Agency | https://www.nisra.gov.uk/statistics/ni-summary-statistics/coronavirus-covid-19-statistics | 1 | 0 | 1 | 0 | 1 |
| **11** | Government of Ireland | https://www.gov.ie/en/ | 0 | 0 | 0 | 0 | 0 |
| **12** | Central Statistics Office Ireland | https://www.cso.ie/en/index.html | 1 | 0 | 1 | 0 | 1 |
| **13** | Public Health Scotland | https://publichealthscotland.scot/ | 16 | 0 | 16 | 0 | 16 |
| **14** | UK COVID-19: testing initiative evaluation programme | https://www.gov.uk/government/collections/covid-19-testing-initiative-evaluation-programme | 7 | 0 | 7 | 0 | 7 |
| | | **TOTAL:** | **52** | **0** | **52** | **0** | **52** |

Grey literature databases search (wellbeing only, reported on the right-hand side of the PRISMA flow diagram in the main text).

| Number of databases | Database name | Website link | Reports sought for retrieval | Reports not retrieved | Reports assessed for eligibility | Reports included | Reports excluded |
|---|---|---|---|---|---|---|---|
| **1** | Opengrey.eu | https://opengrey.eu | 0 | 0 | 0 | 0 | 0 |
| **2** | WHO | https://www.who.int/en/ | 0 | 0 | 0 | 0 | 0 |
| **3** | NTIS (US Department of Commerce) | https://www.ntis.gov | 0 | 0 | 0 | 0 | 0 |
| **4** | WorldCat | https://www.worldcat.org | 8 | 0 | 8 | 0 | 8 |
| **5** | Agency for Healthcare Research and Quality | https://www.ahrq.gov | 0 | 0 | 0 | 0 | 0 |
| | | **TOTAL:** | **8** | **0** | **8** | **0** | **8** |

Studies already identified in other searches are not included here, so as not to double count them.

Google search (wellbeing only, reported on the right-hand side of the PRISMA flow diagram in the main text).

| N of websites | Author | Website link | Reports sought for retrieval | Reports not retrieved | Reports assessed for eligibility | Reports included | Reports excluded |
|---|---|---|---|---|---|---|---|
| **1** | Bahji et al. (2021) | https://www.ncbi.nlm.nih.gov/pmc/articles/PMC8652932/ | * | * | * | * | * |
| **2** | WHO (2020) | Microsoft Word - Mental health considerations 2020-02-14e_en_19MARCH2020_marissa.docx (who.int) | 1 | 0 | 1 | 0 | 1 |
| **3** | Newbigging (2020) | What is the impact of self-isolation and quarantining on our mental health? - University of Birmingham | 1 | 0 | 1 | 0 | 1 |
| **4** | SPI-B (2020) | S0759_SPI-B__The_impact_of_financial_and_other_targeted_support_on_rates_of_self-isolation_or_quarantine_.pdf (publishing.service.gov.uk) | 1 | 0 | 1 | 0 | 1 |
| **5** | Parker et al. (2022) | JMIR Formative Research - A Brief, Daily, Online Mental Health and Well-being Intervention for University Staff During the COVID-19 Pandemic: Program Description and Outcomes Using a Mixed Methods Design | 1 | 0 | 1 | 0 | 1 |
| **6** | Brog et al. (2021) | An internet-based self-help intervention for people with psychological distress due to COVID-19: study protocol for a randomized controlled trial \| Trials \| Full Text (biomedcentral.com) | 1 | 0 | 1 | 0 | 1 |
| **7** | Pietrabissa & Simpson (2020) | Frontiers \| Psychological Consequences of Social Isolation During COVID-19 Outbreak (frontiersin.org) | 1 | 0 | 1 | 0 | 1 |
| **8** | Dagnino et al. (2020) | Frontiers \| Psychological Effects of Social Isolation Due to Quarantine in Chile: An Exploratory Study (frontiersin.org) | 1 | 0 | 1 | 0 | 1 |
| **9** | Wang et al. (2021) | https://www.nature.com/articles/s41380-021-01019-y | 1 | 0 | 1 | 0 | 1 |
| **10** | Young Minds (2020) | Looking After Your Mental Health While Self-isolating \| YoungMinds | 1 | 0 | 1 | 0 | 1 |
| **11** | NHS (nd) | Mental wellbeing while staying at home - Every Mind Matters - NHS (www.nhs.uk) | 1 | 0 | 1 | 0 | 1 |
| **12** | Williams et al. (2021) | Interventions to reduce social isolation and loneliness during COVID-19 physical distancing measures: A rapid systematic review \| PLOS ONE | 1 | 0 | 1 | 0 | 1 |
| **13** | Henssler et al. (2021) | Mental health effects of infection containment strategies: quarantine and isolation—a systematic review and meta-analysis \| SpringerLink | * | * | * | * | * |

| N of websites | Author | Website link | Reports sought for retrieval | Reports not retrieved | Reports assessed for eligibility | Reports included | Reports excluded |
|---|---|---|---|---|---|---|---|
| **14** | Soklaridis et al. (2020) | Mental health interventions and supports during COVID- 19 and other medical pandemics: A rapid systematic review of the evidence - ScienceDirect | 1 | 0 | 1 | 0 | 1 |
| **15** | Schwartz et al. (2021) | COVID-19 and Student Well-Being: Stress and Mental Health during Return-to-School - Kelly Dean Schwartz, Deinera Exner-Cortens, Carly A. McMorris, Erica Makarenko, Paul Arnold, Marisa Van Bavel, Sarah Williams, Rachel Canfield, 2021 (sagepub.com) | 1 | 0 | 1 | 0 | 1 |
| **16** | Hossain et al. (2020) | Mental health outcomes of quarantine and isolation for infection prevention: a systematic umbrella review of the global evidence (e-epih.org) | * | * | * | * | * |
| **17** | UK Government (2022) | COVID-19 Response: Living with COVID-19 - GOV.UK (www.gov.uk) | 1 | 0 | 1 | 0 | 1 |
| **18** | Bunn (2021) | Mental health and well-being in the context of COVID-19 - POST (parliament.uk) | 1 | 0 | 1 | 0 | 1 |
| **19** | CDC (nd) | SARS \| Guidance \| Interventions for Community Containment \| CDC | 1 | 0 | 1 | 0 | 1 |
| **20** | Scottish Government (2021) | Key findings - Compliance with self-isolation and quarantine measures: literature review - gov.scot (www.gov.scot) | 1 | 0 | 1 | 0 | 1 |
| **21** | Bauerle et al. (2020) | e-mental health intervention to support burdened people in times of the COVID-19 pandemic: CoPE It \| Journal of Public Health \| Oxford Academic (oup.com) | 1 | 0 | 1 | 0 | 1 |
| **22** | Suman et al. (2022) | The acceptability of a self-guided psychological intervention for patients with COVID-19 in isolation and quarantine \| International Journal Of Community Medicine And Public Health (ijcmph.com) | 1 | 0 | 1 | 0 | 1 |
| **23** | CAMH (nd) | Quarantine & isolation \| CAMH | 1 | 0 | 1 | 0 | 1 |
| **24** | Cherry (2020) | Protect Your Mental Health During Quarantine (verywellmind.com) | 1 | 0 | 1 | 0 | 1 |
| **25** | Jurblum et al. (2020) | RACGP - Psychological consequences of social isolation and quarantine | 1 | 0 | 1 | 0 | 1 |
| **26** | Bahji et al. (2021) | (18) (PDF) Strategies to aid self-isolation and quarantine for individuals with severe and persistent mental illness during the COVID-19 pandemic: A systematic review (researchgate.net) | * | * | * | * | * |
| **27** | Ankit et al. (2020) | Impact on mental health by “Living in Isolation and Quaranti... : Journal of Family Medicine and Primary Care (lww.com) | 1 | 0 | 1 | 0 | 1 |

| N of websites | Author | Website link | Reports sought for retrieval | Reports not retrieved | Reports assessed for eligibility | Reports included | Reports excluded |
|---|---|---|---|---|---|---|---|
| **28** | BMA (nd) | bma-the-impact-of-covid-19-on-mental-health-in-england.pdf | 1 | 0 | 1 | 0 | 1 |
| **29** | Soklaridis et al. (2020) | Mental health interventions and supports during COVID- 19 and other medical pandemics _ A rapid systematic review of the evidence (careknowledge.com) | 1 | 0 | 1 | 0 | 1 |
| **30** | Reagu et al. (2020) | Psychological impact of the COVID-19 pandemic within institutional quarantine and isolation centres and its sociodemographic correlates in Qatar: a cross-sectional study \| BMJ Open | * | * | * | * | * |
| **31** | Positive Mind Works (nd) | Self-Isolation Program \| Heading into Quarantine? Let us help you through (positivemindworks.co) | 1 | 0 | 1 | 0 | 1 |
| **32** | Public Health Agency Belfast (2021) | How to self-isolate in a shared house if you or someone you live with has coronavirus \| HSC Public Health Agency (hscni.net) | 1 | 0 | 1 | 0 | 1 |
| **33** | European Centre for Disease Prevention and Control (nd) | Guidance on quarantine of close contacts to COVID-19 cases and isolation of COVID-19 cases, 7 January 2022 (europa.eu) | 1 | 0 | 1 | 0 | 1 |
| **34** | National Elf Service (2020) | Quarantine: infection prevention, but at what cost for mental health? - National Elf Service | 1 | 0 | 1 | 0 | 1 |
| **35** | Eraso et al. (2021) | IJERPH \| Free Full-Text \| Self-Isolation and Quarantine during the UK's First Wave of COVID-19. A Mixed-Methods Study of Non-Adherence (mdpi.com) | * | * | * | * | * |
| **36** | Lingnan University (2022) | LU designs self-assessment test to improve the psychological wellbeing of people in quarantine - Press Releases - Media - Lingnan University (ln.edu.hk) | 1 | 0 | 1 | 0 | 1 |
| **37** | Imperial College (2020) | Imperial-College-COVID19-NPI-modelling-16-03-2020.pdf | 1 | 0 | 1 | 0 | 1 |
| **38** | Center for the Study of Traumatic Stress (nd) | Psychological Effects of Quarantine (cstsonline.org) | 1 | 0 | 1 | 0 | 1 |
| **39** | Hossain et al. (2020) | Mental health outcomes of quarantine and isolation for infection prevention: A systematic umbrella review of the global evidence - Abstract - Europe PMC | * | * | * | * | * |
| **40** | Nuffield Trust (2021) | To solitude: Learning from other countries on how to improve compliance with self-isolation \| Nuffield Trust | 1 | 0 | 1 | 0 | 1 |
| **41** | Australian Government (2022) | Pandemic (Quarantine Isolation and Testing Order) 2022 (No. 11).pdf (health.vic.gov.au) | 1 | 0 | 1 | 0 | 1 |

| N of websites | Author | Website link | Reports sought for retrieval | Reports not retrieved | Reports assessed for eligibility | Reports included | Reports excluded |
|---|---|---|---|---|---|---|---|
| **42** | Sanders (2020) | COVID-19, social isolation and loneliness \| Iriss | 1 | 0 | 1 | 0 | 1 |
| **43** | Kumar et al. (2020) | Full article: COVID 19 and its mental health consequences (tandfonline.com) | 1 | 0 | 1 | 0 | 1 |
| **44** | Australian Government (2022) | nchrac-report-Mental-health-impacts-of-quarantine-and-self-isolation0620.pdf (nhmrc.gov.au) | 1 | 0 | 1 | 0 | 1 |
| **45** | Panchal et al (2023) | The Implications of COVID-19 for Mental Health and Substance Use \| KFF | 1 | 0 | 1 | 0 | 1 |
| **46** | Jassim et al. (2021) | Psychological impact of COVID-19, isolation and quarantine \| NDT (dovepress.com) | * | * | * | * | * |
| **47** | NIHR (2021) | Lonely young people have an increased risk of mental health problems years later: research suggests lockdown could have a long term effect (nihr.ac.uk) | 1 | 0 | 1 | 0 | 1 |
| **48** | Smith (2022) | Learning lessons from the UK's self-isolation programme - King's College London (kcl.ac.uk) | 1 | 0 | 1 | 0 | 1 |
| **49** | Ho et al. (2020) | Ho-et-al.-2020-Mental-health-strategies-during-Covid-19.pdf (lancsvrn.co.uk) | 1 | 0 | 1 | 0 | 1 |
| **50** | Organisation for Economic Co-operation and Development (2020) | COVID-19: Protecting people and societies (oecd.org) | 1 | 0 | 1 | 0 | 1 |
| **51** | Nuffield (2020) | Ethical-considerations-in-responding-to-the-COVID-19-pandemic.pdf (nuffieldbioethics.org) | 1 | 0 | 1 | 0 | 1 |
| **52** | Local Government Association (nd) | Public mental health and wellbeing and COVID-19 \| Local Government Association | 1 | 0 | 1 | 0 | 1 |
| **53** | Queen's University Belfast (nd) | Supporting Children in Isolation \| Coronavirus (COVID-19) \| Queen's University Belfast (qub.ac.uk) | 1 | 0 | 1 | 0 | 1 |
| **54** | All India Occupational Therapists Federation (nd) | Role of Occupational Therapist in COVID-19 (wfot.org) | 1 | 0 | 1 | 0 | 1 |
| **55** | AMA (nd) | Ethical Use of Quarantine & Isolation \| ama-coe (ama-assn.org) | 1 | 0 | 1 | 0 | 1 |
| **56** | Australian government (2020) | National Mental Health and Wellbeing Pandemic Response Plan (mentalhealthcommission.gov.au) | 1 | 0 | 1 | 0 | 1 |
| **57** | Okabe-Miyamoto et al. (2020) | Social Connection and Well-Being during COVID-19 \| The World Happiness Report | 1 | 0 | 1 | 0 | 1 |
| **58** | Hester (2020) | The toll of COVID-19 and quarantine on college students' mental health (contemporarypediatrics.com) | 1 | 0 | 1 | 0 | 1 |

| N of websites | Author | Website link | Reports sought for retrieval | Reports not retrieved | Reports assessed for eligibility | Reports included | Reports excluded |
|---|---|---|---|---|---|---|---|
| **59** | Save the Children (2021) | Integrated Response Framework for Isolation and Quarantine as Non-Pharmaceutical Interventions Against COVID-19 (ready-initiative.org) | 1 | 0 | 1 | 0 | 1 |
| **60** | Louisiana Department of Health (2022) | LDH_COVID-Contact-Tracing-in-Schools.pdf (la.gov) | 1 | 0 | 1 | 0 | 1 |
| **61** | Unicef (2020) | Briefing note on addressing mental health and psycosocial aspects of COVID-19 outbreak.pdf (unicef.org) | 1 | 0 | 1 | 0 | 1 |
| **62** | Physiopedia (nd) | Mental Health Stress and Resilience in Times of COVID-19 - Physiopedia (physio-pedia.com) | 1 | 0 | 1 | 0 | 1 |
| **63** | Zweig et al. (2021) | Ensuring Rights while Protecting Health: The Importance of Using a Human Rights Approach in Implementing Public Health Responses to COVID-19 – Health and Human Rights Journal (hhrjournal.org) | * | * | * | * | * |
| **64** | Fuchs et al. (2021) | Assessment of a Hotel-Based COVID-19 Isolation and Quarantine Strategy for Persons Experiencing Homelessness \| Infectious Diseases \| JAMA Network Open \| JAMA Network | * | * | * | * | * |
| **65** | Lockhart et al. (2020) | CYP-MH-Restoration-Recovery-Planning-Literature-Summary-v.2.0.docx (live.com) | 1 | 0 | 1 | 0 | 1 |
| **66** | California Department of Public Health (2020) | https://www.cdph.ca.gov/Programs/CID/DCDC/Pages/COVID-19/Guidance-on-Isolation-and-Quarantine-for-COVID-19-Contact-Tracing-7-30-2020.aspx | 1 | 0 | 1 | 0 | 1 |
| **67** | Lewis & Clark (2020) | COVID-19 Isolation and Quarantine Plan for the Lewis & Clark Community • Health Promotion and Wellness • Lewis & Clark (lclark.edu) | 1 | 0 | 1 | 0 | 1 |
| **68** | Cal Poly (nd) | COVID-19 - Campus Health & Wellbeing - Cal Poly, San Luis Obispo | 1 | 0 | 1 | 0 | 1 |
| **69** | Newman University (2020) | Covid-19-Resilience-Document-May-2022-Final.pdf (newman.ac.uk) | 1 | 0 | 1 | 0 | 1 |
| **70** | Vermont Department of Health (nd) | COVID-19 Symptoms & Treatment \| Vermont Department of Health (healthvermont.gov) | 1 | 0 | 1 | 0 | 1 |
| **71** | Arizona Department of Health Services (2022) | ADHS 'Release from Isolation and Quarantine' Guidance (azdhs.gov) | 1 | 0 | 1 | 0 | 1 |
| **72** | FutureLearn (nd) | How to Survive Self-Isolation - Blog - FutureLearn | 1 | 0 | 1 | 0 | 1 |
| **73** | Government of Canada (nd) | COVID-19: Prevention and risks - Canada.ca | 1 | 0 | 1 | 0 | 1 |

| N of websites | Author | Website link | Reports sought for retrieval | Reports not retrieved | Reports assessed for eligibility | Reports included | Reports excluded |
|---|---|---|---|---|---|---|---|
| **74** | Vanderbilt University (nd) | Support During Quarantine and Isolation \| Student Care Network \| Vanderbilt University | 1 | 0 | 1 | 0 | 1 |
| **75** | New Zealand Government (2020) | COVID-19 Preliminary Psychosocial and Mental Wellbeing Recovery Plan (health.govt.nz) | 1 | 0 | 1 | 0 | 1 |
| **76** | Diamond et al. (2020) | Coronavirus disease 2019: achieving good mental health during social isolation. — Department of Psychiatry (ox.ac.uk) | * | * | * | * | * |
| **77** | USA Government (nd) | What is the difference between isolation and quarantine? \| HHS.gov | 1 | 0 | 1 | 0 | 1 |
| **78** | US Department of Education (nd) | Supporting Students During the COVID-19 Pandemic: Maximizing In-Person Learning and Implementing Effective Practices for Students in Quarantine and Isolation \| U.S. Department of Education | 1 | 0 | 1 | 0 | 1 |
| **79** | Norwegian Institue of Public Health (nd) | Coronavirus disease - NIPH (fhi.no) | 1 | 0 | 1 | 0 | 1 |
| **80** | New York (2023) | Monroe County, NY - COVID-19 Resources | 1 | 0 | 1 | 0 | 1 |
| **81** | Office for Students (2020) | Supporting student mental health - Office for Students | 1 | 0 | 1 | 0 | 1 |
| **82** | Voyae Well (nd) | Voyage Well \| Virgin Voyages | 1 | 1 | 0 | 0 | 0 |
| **83** | Unknown | Revised_Travel_protocol_revised_2nd_April_2022.pdf (ncdc.gov.ng) | 1 | 1 | 0 | 0 | 0 |
| **84** | Wikipedia (nd) | COVID-19 pandemic - Wikipedia | 1 | 0 | 1 | 0 | 1 |
| **85** | Tulane University (2020) | Understanding the Effects of Social Isolation on Mental Health (tulane.edu) | 1 | 0 | 1 | 0 | 1 |
| **86** | Australian Government (2023) | CHOICE travel insurance buying guide \| Smartraveller | 1 | 0 | 1 | 0 | 1 |
| **87** | Czech Government (2022) | Measures adopted by the Czech Government against the coronavirus \| Government of the Czech Republic (vlada.cz) | 1 | 0 | 1 | 0 | 1 |
| **88** | State of New Jersey (2022) | https://www.cityofsummit.org/660/COVID-19 | 1 | 0 | 1 | 0 | 1 |
| **89** | Cayman Islands Government (2023) | COVID-19 Frequently Asked Questions (exploregov.ky) | 1 | 0 | 1 | 0 | 1 |
| **90** | WebMD (2023) | Review Supports Continued Mask-wearing in Health Care Visits (webmd.com) | 1 | 0 | 1 | 0 | 1 |

| N of websites | Author | Website link | Reports sought for retrieval | Reports not retrieved | Reports assessed for eligibility | Reports included | Reports excluded |
|---|---|---|---|---|---|---|---|
| **91** | Johns Hopkins (2020) | Coronavirus, Social and Physical Distancing and Self-Quarantine \| Johns Hopkins Medicine | 1 | 0 | 1 | 0 | 1 |
| **92** | Monson Town Government (2022) | Coronavirus / COVID19 Information \| Town of Monson MA (monson-ma.gov) | 1 | 0 | 1 | 0 | 1 |
| **93** | na | Click Allow (solidcaptcha.lm.r.appspot.com) | 1 | 1 | 1 | 0 | 0 |
| **94** | US Department of Health and Human Services (nd) | SAMHSA's National Helpline \| SAMHSA | 1 | 0 | 1 | 0 | 1 |
| **95** | Moustafa (nd) | Mental Health Effects of COVID-19 - Google Books | 1 | 0 | 1 | 0 | 1 |
| **96** | Viditch (nd) | Germs at Bay: Politics, Public Health, and American Quarantine - Charles Vidich - Google Books | 1 | 0 | 1 | 0 | 1 |
| **97** | Freeman (nd) | The Ethics of Public Health, Volumes I and II - Google Books | 1 | 0 | 1 | 0 | 1 |
| **98** | Bahji et al. (2021) | Strategies to aid self-isolation and quarantine for individuals with severe and persistent mental illness during the COVID-19 pandemic: A systematic review \| Psychiatric Research and Clinical Practice (psychiatryonline.org) | * | * | * | * | * |
| **99** | Hossain et al. (2020) | Mental health outcomes of quarantine and isolation for infection prevention: a systematic umbrella review of the global evidence - PMC (nih.gov) | * | * | * | * | * |
| **100** | Jain et al. (2020) | Impact on mental health by "Living in Isolation and Quarantine" during COVID-19 pandemic - PMC (nih.gov) | * | * | * | * | * |
| | | **TOTAL:** | **86** | **3** | **83** | **0** | **83** |

* Study had already been identified in a previous search (database or other grey literature) and therefore was not screened again.

**Appendix 3: Deviation from the protocol published on PROSPERO**

*Additional exclusion criteria and clarification of the application of existing criteria*

The initial screening against the criteria specified in the protocol produced 164 citations for inclusion, deemed too many for effective synthesis. To reduce the number of studies, we examined heterogeneity in the study populations. We excluded studies that examined populations of children and students (N=23), health care workers (N=23), managed isolation (for example, hotels and self-isolation due to travel because usually this was in a hotel or other institution) N=82. Note that excluding isolation in a hospital setting was already specified in the pre-registration.

We also excluded studies, or specific findings from studies, where the wellbeing outcome was not specified in the search terms, which could lead to non-identification of all studies related to this outcome. Two quantitative studies were excluded because the outcome was intimate partner violence.

During the pilot training set, many studies were identified where the definition of isolation and quarantine did not match our definition for the purpose of this review. For example, isolation could refer to self-imposed isolation due to vulnerability, or lockdown measures. Therefore, additional guidance for reviewers was added:

- For full-text screening, the abstract must, in principle, have captured self-isolation as defined for this study.
- Once full text-screening was complete, if it remained unclear whether self-isolation matched our definition, we contacted the authors. If it remained unclear, we excluded the study from the review.

We also introduced additional criteria to aid screening/searching:

- For any of the excluded characteristics, we allowed up to 5% of participants to have that characteristic before excluding the study.
- Grey literature was only included if it investigated the effectiveness of an intervention to ensure only the most rigorous non-peer-reviewed studies were included and because of the dearth of peer-reviewed data on this specific topic.

*Other deviations*

We included a search of the Embase database, which was not specified in the protocol. The protocol specified the use of NIH quality assessment tools. Following consultation of the Cochrane handbook, we decided instead to use the Risk Of Bias In Non-randomized Studies (ROBINS) for exposure and intervention studies. We therefore did not collect data on funding information as planned, as this is not required for the ROBINS assessment. We used the CASP assessment tool for the qualitative studies because the Cochrane recommended ROBINS tools do not currently include a qualitative assessment.

**Appendix 4: Quality assessment tools**

Risk of bias assessments were undertaken by one reviewer, and each was discussed with another reviewer for consistency. We used ROBINS-E for the assessment of non-randomised studies of exposure and ROBINS-I for the assessment of non-randomized studies of interventions,[14,15] guided by the *Cochrane Handbook for Systematic Reviews of Interventions* for assessing risk of bias of different types of non-randomized studies.[10]

ROBINS-E and ROBINS-I require the pre-identification of potentially significant confounding domains. We specified gender and age, which were applied to all studies. Using the ROBINS algorithm, studies that do not achieve this a priori confounding considerations are not assessed further, due to their substantial risk of bias. Nevertheless, we evaluated all risk of bias domains for all included studies to provide a comprehensive perspective on bias risk within each domain across the entire set of studies. In cases where studies did not meet the a priori criterion, they were assigned an overall bias risk rating of 'very high.'

ROBINS assessments are specific to a reported result rather than a study. Consequently, studies that reported a result for aim 1 and 2 received two risk of bias scores.

For consistency of nomenclature, ratings for the ROBINS-I were converted to use the same terminology as the ROBINS-E: ROBINS-I “low risk of bias” = low risk of bias; ROBINS-I “moderate risk of bias” = some concerns; ROBINS-I “serious risk of bias” = high risk of bias; ROBINS-I “critical risk of bias” = very high risk of bias). No amendments were made to the algorithms used to determine the risk of bias rating.

We used the Critical Appraisal Skills Programme (CASP) checklist for qualitative studies,[16] but reworded the item ‘how valuable is the research?’ to ‘do the authors discuss the value of the research in terms of implications and contribution to literature?’ to allow yes/no responses in line with the other items and to give each study an overall quality score percentage. Scores were out of ten, reported as a percentage, with higher scores indicating better quality.

**Appendix 5: Synthesis**

Quantitative data were synthesised narratively, following SWiM guidelines.[17] No meta-analysis was planned due to expected heterogeneity in study design, outcomes, and associated factors. Studies were synthesised within each research aim separately and grouped by psychological wellbeing outcome to reduce heterogeneity. Studies reporting general psychological symptom scores (i.e., not disorder-specific) or subjectively reported mental health (e.g., 'compared to before the COVID-19 pandemic, how would you say your mental health is now?' [25]) were grouped together as *general psychological symptoms*. To be included in the synthesis, each wellbeing outcome (aim 1) and each factor (aim 2) had to be investigated in at least two studies. If removing one study from the synthesised table for this reason left only one study for another outcome/factor, that study was also removed from the table.

For aim 1, we compared outcomes for those in self-isolation with those not self-isolating. For aim 2, we also grouped studies by factor (isolation, demographic, COVID-19, and mental/physical health characteristics) and associations between factors and self-isolation were reported. These groupings were not defined a priori and emerged from the data during tabulation. For aim 3, we grouped studies by intervention type and compared pre- and post-intervention scores in the intervention group when there was no control group, and compared the intervention group to the control group where one was reported. Studies were synthesised using tabulation and vote counting based on the direction of effect. The number of studies, the consistency of effects across studies, and the risk of bias across studies were used to assess the certainty of synthesised findings.

Qualitative data were synthesised using meta-ethnography, following eMERGe guidelines.[18] Subthemes were developed by psychological wellbeing outcome and for aim 2, subthemes were developed by factor. No qualitative studies explored interventions (aim 3). Qualitative findings were synthesised using reciprocal translation (to understand one study's findings in terms of another) and refutational translation (to explore differences between studies and differences identified in quantitative findings).

### Appendix 6: Data extraction tables

*Table 1. Isolation characteristics*

| Citation | Reason for isolation/quarantine (infected, suspected to be infected, close contact) | Location (make this home if stated, assumed to be home if not); duration of isolation/quarantine; lockdown context | Assessment points; amount of time between isolation/quarantine and data collection |
|---|---|---|---|
| **Aaltonen et al., 2022** [19] | Infected, close contact. | Home; the mean quarantine duration for the quarantined sample was 10.8 days (SD=3.4); not reported. | Baseline = cases were telephone interviewed about at 1 week (baseline) since the onset of the quarantine and the controls within 10 days since PCR testing.<br><br>Follow up = interviews were conducted mean 15.6 [SD 2.4] days since the onset of quarantine and mean 4.9 [SD 3.5] days since the expiration. |
| **Abir et al., 2021** [20] | Infected, close contact. | Assumed to be home; not reported; mandatory lockdown was in place in parts of Bangladesh, before the vaccine rollout. | 'currently in self-quarantine since past 7 days' |
| **Aloba & Opakunle, 2021** [21] | Infected. | Home; mean duration = 8.79 days (SD=2.71) range=1-14; not reported. | Participants given a code for the online survey at the treatment centre, days before completing survey not reported |
| **Aslaner et al., 2022** [22] | Infected, suspected to be infected, close contact. | Home; not reported; not reported. | Not reported |
| **Bonsaksen et al., 2020** [23] | Infected, close contact. | Assumed to be home; not reported; Norwegian authorities imposed a lockdown of society on 12 March 2020. While restrictions were lifted gradually during the following months, people were encouraged to shelter in place. Kindergartens, schools, and universities were closed, as were non-vital businesses requiring physical proximity, and cultural events and travels were cancelled. | Not reported: 'have been in quarantine/isolation' |
| **Chakeri et al., 2020** [24] | Infected. | Home; not reported; not reported. | Pre-intervention = the start of isolation (day 0)<br><br>Follow up = day 21 |

| Citation | Reason for isolation/quarantine (infected, suspected to be infected, close contact) | Location (make this home if stated, assumed to be home if not); duration of isolation/quarantine; lockdown context | Assessment points; amount of time between isolation/quarantine and data collection |
|---|---|---|---|
| **Daly et al., 2021** [25] | Infected, suspected to be infected, close contact. | Assumed to be home; 10-14 days; Data collection was approximately one month after the initial peak of the COVID-19 pandemic in Canada and coincides with the time during which many jurisdictions began their initial phases of "re-opening", following approximately two months of widespread restrictions. | 1 assessment point; retrospectively reported 'since the start of the pandemic', which was ~6-8 weeks before data collection. |
| **Domenghino et al., 2022** [26] | Infected. | For the whole sample: 92.8% at home, 2.1% at someone else's home, 4.9% in hospital, 0.2% at a social institution, 0.1% in a hotel, 2.1% 'other', 1.3% missing data. Additionally, 55 participants isolated in more than one place, in most cases (N=41) first at hospital and then at home; Retrospectively recruited group: median number of days in isolation 10.0, range 2.0-25.0 (N=276, 25%, had missing data for this), prospective group: not reported; not reported. | Retrospective group completed baseline measures a median of 7.2 months after diagnosis; prospective group completed baseline measures 'upon or shortly after diagnosis'<br><br>Both groups completed follow-up measures two weeks and four weeks after baseline |
| **Flores-Torres et al., 2021** [27] | Suspected to be infected , close contact. | Assumed to be home; not reported; during national stay-at-home directives. | Self-isolation in the past week |
| **Gok, 2022** [28] | Infected. | Home; not reported; not reported. | Not reported |
| **Havlioglu et al., 2022** [29] | Infected. | Home; 14 days; 'Some of the measures … not leaving the house except for necessities, closing schools, and telecommuting in public offices'. | Not reported |
| **Isherwood et al., 2022** [30] | Close contact. | Home; 10 or 14 days; not reported. | Not reported |
| **Jagadeesan et al., 2022** [31] | Infected. | Home; 15 days; not reported. | Pre-intervention: start of isolation (day 0)<br><br>Post-intervention: end of isolation (day 16) |
| **Jang et al., 2022** [32] | Infected, close contact. | Assumed to be at home; not reported; not reported. | Not reported |
| **Jesmi et al., 2021** [33] | Infected. | Assumed to be at home; not reported; not reported. | Not reported |

| Citation | Reason for isolation/quarantine (infected, suspected to be infected, close contact) | Location (make this home if stated, assumed to be home if not); duration of isolation/quarantine; lockdown context | Assessment points; amount of time between isolation/quarantine and data collection |
|---|---|---|---|
| **Joisten et al, 2022** [34] | Infected, close contact. | Home; mean = 11.8 days, SD = 4.6 days; not reported. | Not reported |
| **Ju et al., 2021** [35] | Infected. | Home vs hotel; two weeks; not reported. | Baseline = discharge day<br><br>Follow up = two weeks after discharge, when isolation complete, days before completing the survey not reported |
| **Kopilas et al., 2021** [36] | Close contact. | Home; two weeks; lockdown and non-lockdown contexts. | During the isolation period |
| **Kowalski et al., 2021** [37] | Infected. | Home; 14 days; not reported. | During isolation (M=12.7 days, SD=4.6) |
| **Lohiniva et al., 2021** [38] | Infected, close contact. | Home; not reported; not reported. | Interviews were all conducted at least 28 days after the household index's PCR confirmation; PCR confirmed cases occurred during February-April 2020 and interviews took place during April – May 2020 |
| **Maric et al., 2022** [39] | Close contact. | Assumed to be at home; not reported; between pandemic peaks, limited restrictions in place and substantial vaccination rates. | During interview; not reported |
| **Mohamed & Yousef, 2021** [40] | Infected. | Home vs hospital; not reported; not reported | Not reported |
| **Navas et al., 2022** [41] | Infected. | Home vs hotel; 14 days; during data collection, Spain was the country with the second highest number of infected people and the third in number of deaths. A number of restrictive measures had been in place for two months. | One assessment point; during isolation treatment |
| **Oginni et al., 2021** [42] | Suspected to be infected. | Assumed to be home; not reported; during national lockdown. | Not reported |
| **Opakunle, 2022** [43] | Infected. | Home; not reported; not reported. | Not reported; Ct values taken from PCR test at presentation to treatment centre |

| Citation | Reason for isolation/quarantine (infected, suspected to be infected, close contact) | Location (make this home if stated, assumed to be home if not); duration of isolation/quarantine; lockdown context | Assessment points; amount of time between isolation/quarantine and data collection |
|---|---|---|---|
| **Paz et al., 2020** [44] | Infected, suspected to be infected. | Assumed to be at home; not reported; not reported | Not reported |
| **Petrocchi et al., 2021** [45] | Infected. | Home vs hospital; not reported; during national lockdown | Quarantine during 22 March to 11 May 2020; 5-7 months before data collection |
| **Pheh et al., 2020** [46] | Not reported. | Home; not reported; during nationwide movement control order (i.e. lockdown). | Baseline (at recruitment into study)<br>Post treatment (same day as intervention)<br>Follow-up (2 weeks after the end of treatment) |
| **Plesea-Condratovici et al., 2022** [47] | Infected. | Home; 14 days; not reported | GP daily phone monitoring during isolation |
| **Rajagopalan et al., 2022** [48] | Infected. | Home; 14 days; not reported | Pre-intervention – start of isolation (day 0)<br>Post-intervention – end of isolation (day 15) |
| **Ripon et al., 2020** [49] | Infected. | Home vs instituition; 14 days; National lockdown had been extended to cover the data collection period | Within 5 days of end of quarantine |
| **Rossi et al., 2020** [50] | Infected, close contact. | Assumed to be home; not reported; during nationwide lockdown 'included travel restrictions, the mandatory closure of schools, nonessential commercial activities and industries. People were asked to stay at home and socially isolate themselves to prevent being infected' and 'the investigated timeframe corresponds to the contagion peak in Italy, according to epidemiological data confirmed by the World Health Organization'. | Currently in quarantine (i.e. self-isolation) |
| **Schluter et al., 2022** [51] | Close contact, suspected to be infected, infected, travel/health. | Assumed to be home; not reported; 1 year after first reported case, 'Selection of countries for inclusion was based on ensuring global continent diversity and … capturing different demographics, health systems and policies, and COVID-19 burdens and response'. | Not reported; within 12 months following the earliest reported COVID-19 case. |

| Citation | Reason for isolation/quarantine (infected, suspected to be infected, close contact) | Location (make this home if stated, assumed to be home if not); duration of isolation/quarantine; lockdown context | Assessment points; amount of time between isolation/quarantine and data collection |
|---|---|---|---|
| **Wessely et al., 2022** [53] | Close contact, infected. | Home; 10-14 days (mean=11.8 days, SD=4.6); not reported. | 1 assessment point; up to 11 months |
| **Verberk et al., 2021** [52] | Close contact. | Home; at least 7 days and 24 hours symptom free; not reported. | 7-15 days after the COVID-19 diagnosis of the index case |
| **Xu et al., 2020** [54] | Close contact. | Home; not reported; not reported. | 1 assessment point, during quarantine |

*Table 2. Quantitative outcomes by research aim*

| Citation | Measures of psychological wellbeing | Impact of self-isolation on wellbeing: Test (relationship analysed) *bold indicates significance *italicised indicates predictor (isolation type)* | Factors associated with wellbeing: Test (relationship analysed) *bold indicates significance *italicised indicates wellbeing outcome* | Intervention outcome on wellbeing Test (relationship analysed) *bold indicates significance *italicised indicates wellbeing outcome* |
|---|---|---|---|---|
| **Aaltonen et al., 2022** [19] | CORE-OM Psychic wellbeing and distress symptom scores and subscales<br><br>Patient Health Questionnaire (PHQ-9; depression symptoms)<br><br>Overall Anxiety Severity and Impairment Scale (OASIS; anxiety symptoms)<br><br>Binarised using established thresholds indicating a clinical diagnosis or severity classification | Multivariate linear regression<br>Overall psychological wellbeing symptom score at baseline (7-10 days after PCR/start of quarantine) in the whole sample (*being quarantined was not significantly associated*)<br><br>Kruskal-Wallis one way ANOVA CORE-OM – Subjective wellbeing<br>CORE-OM – Problems/symptoms<br>**CORE-OM – Life functioning (*all quarantined/isolating > controls*)**<br>CORE-OM – Risk/harm<br>Chi-square test (unadjusted)<br>CORE-OM – Distress<br><br>**only adjusted estimates are reported in the summary table (Aim 1 ROB relates to the adjusted analysis)* | Chi square tests (unadjusted) comparing reason for quarantine (infected vs close contacts) at 2-week follow-up<br>*Overall psychological wellbeing clinical outcome*<br>***Depression clinical outcome* (infected > close contac**t)<br>*Anxiety clinical outcome*<br><br>Kruskal-Wallis one way ANOVA (unadjusted) comparing reason for quarantine (infected vs close contacts) at follow-up)<br>CORE-OM – Total score<br>CORE-OM – Subjective wellbeing<br>CORE-OM – Problems/symptoms<br>CORE-OM – Life functioning<br>CORE-OM – Risk/harm<br>Chi-square test (unadjusted)<br>CORE-OM – Distress | Not assessed |
| **Abir et al., 2021** [20] | Impact of Event Scale Revised (IES-R, psychological impact)<br><br>Binarised: mild to severe vs none | Logistic regression (adjusted odds ratio)<br><br>**Psychological impact (*quarantine > no quarantine*)** | Not reported | Not reported |
| **Aloba & Opakunle, 2021** [21] | Wellbeing outcome: Insomnia Severity Index (ISI)<br><br>Predictors:<br>Generalized Anxiety Disorder-7 scale (GAD-7; anxiety symptoms) | Not reported | Multivariate linear regression:<br><br>*Insomnia:*<br>Age<br>Viral load<br>Anxiety | Not reported |

| | | | | |
|---|---|---|---|---|
| | Patient Health Questionnaire (PHQ-8; depression symptoms)<br>Suicidal ideation (item 9 of the PHQ-9)<br>Brief Symptoms Rating Scale (BSRS-5; general psychological symptoms). | | Depression<br>**General psychological symptoms (positive association)**<br>**Suicidal ideation (positive association)**<br>**Duration of isolation (negative association)** | |
| **Aslaner et al., 2022** [22] | Coronavirus Anxiety Scale (CAS)<br><br>Death Anxiety Scale (DAS), how categorised into y/n is not reported | Not reported | Logistic regression (adjusted odds ratio)<br>*Death anxiety*<br>**Gender (male > female)**<br>**Positive PCR (yes > no)**<br>**Presence of symptoms (no > yes)** | Not reported |
| **Bonsaksen et al., 2020** [23] | PTSD Checklist (PCL-5; PTSD) to measure symptoms, binarised using DSM-5 diagnostic guidelenes to categorise y/n | Logistic regression (adjusted odds ratios)<br><br>**PTSD (*been in quarantine/isolation > not been in quarantine/isolation*)** | Not reported | Not reported |
| **Chakeri et al., 2020** [24] | The Iranian version of the Spielberger Anxiety Inventory (SAI; anxiety symptoms) | Not reported | Not reported | Independent t-test<br><br>*Anxiety symptoms following the intervention* **(pre > post)**<br><br>*Anxiety symptoms at follow up* **(Controls > intervention)** |
| **Daly et al., 2021** [25] | Subjective report: 'compared to before the COVID-19 pandemic, how would you say your mental health is now?' 5 point likert scale, slightly and significantly worse were collapsed into outcome 'worse mental health'<br><br>Suicidal thoughts and self harm, separate items: 'as a result of the COVID-19 pandemic, in the previous two weeks, have you experienced either suicidal thoughts/feelings' or 'deliberately hurt yourself' | Chi-square test:<br>**Rates of subjective worse mental health (*isolated for any reason (EXCLUDING isolate for travel) > those who had not isolated*)** | Not reported | Not reported |
| **Flores-Torres et al., 2021** [27] | The Spanish version of the 7-item Center for Epidemiologic Studies Depression scale (CESD-7), categorised to be clinically significant y/n | Logistic regressions (adjusted odds ratio)<br><br>**depression (*self-isolated in past week > not self-isolated in past week*)**<br>anxiety | Not reported | Not reported |

| | | | | |
|---|---|---|---|---|
| | The Spanish version of the Generalized Anxiety Disorder-7 scale (GAD-7), categorised to be clinically significant y/n<br><br>Subjective item: has worry or stress related to the coronavirus had a negative impact on your mental health? (binarised y/n) | mental health impact | | |
| **Havlioglu et al., 2022** [29] | The Turkish version of the Padua Inventory-Revised (Padua; Obsessive-compulsive symptoms) | Not reported | Individual ANOVAs and post hoc Tukey tests for pairwise group differences<br><br>*Obsessive compulsive symptoms:*<br>Gender<br>Age<br>**Education (degree > high school/elementary school)**<br>SES<br>**Comorbid disease (yes > no)**<br>Time of year (season)<br>**Psychiatric illness (yes > no)** | Not reported |
| **Isherwood et al., 2022** [30] | Subjective self-reported mental health difficulties (one item) and loneliness (one item) | Not reported | Logistic regression (adjusted odds ratios)<br><br>*Mental health difficulties*<br>**Age (younger > older)**<br>**Gender (female > male)**<br>Living alone<br><br>*Loneliness*<br>**Age (younger > older)**<br>Gender<br>**Living alone (no > yes)** | Not reported |
| **Jagadeesan et al., 2022** [31] | Depression Anxiety and Stress Scale-21 (DASS-21; total symptom score, depression score, anxiety score, stress score)<br><br>Pittsburgh Sleep Quality Index (PSQI; sleep quality) | Not reported | Not reported | Paired t-test (unadjusted)<br><br>***Total symptoms* (baseline > follow up)**<br>***Depression* (baseline > follow up)**<br>***Anxiety* (baseline > follow up)** |

| | | | | |
|---|---|---|---|---|
| | WHO Quality of Life (WHOQ<br>OL-BREF; psychological subscale) | | | ***Stress* (baseline > follow up)**<br>***Insomnia* (baseline > follow up)**<br>***Quality of life* (follow up > baseline)** |
| **Jang et al., 2022 [32]** | Patient Health Questionnaire-2 (PHQ-2; depression symptoms) binarised 'subject had depressive symptoms' y/n | Not reported | One logistic regression model (adjusted odds ratios)<br><br>*Depression symptoms:*<br>**Age (<40y > 40-64y and 65y+)**<br>Gender<br>**Education (<middle school > college)**<br>**Income (highest > all lower categories)**<br>**Changes in daily life due to COVID-19 (no > yes)**<br>Employment<br>**Poor physical health (yes > no)**<br>Married<br>Living alone<br>**Assistance with isolation (no > yes)**<br>**Counselling for depression (yes > no)** | Not reported |
| **Joisten et al, 2022 [34]** | Psychological stress<br><br>Five items adapted from the COVID-19 Snapshot Monitoring (COSMO) questionnaire from the University of Erfurt<br><br>In detail, item 1 from the generalized anxiety disorder (GAD-7) items 6, 8 and 14 from the Allgemeine Depressionsskala (ADS) and item 19 from the IES-R24 (impact of event scale). *<br><br>**The review authors retrieved this assessment from the study protocol* | Not reported | Test not reported.<br><br>**Infected > close contact** | Not reported |

| | | | | |
|---|---|---|---|---|
| **Ju et al., 2021** [35] | Chinese version of the 9-item Patient Health Questionnaire (PHQ-9; depression symptoms)<br><br>Chinese version of the 7-item Generalized Anxiety Disorder scale (GAD-7; anxiety symptoms) | Not reported | General linear model with repeated measures to examine the effects of time by isolation type on depression, anxiety. Covariates (demographics, disease-related, counselling during isolation) did not predict the outcome in the mulitivariate model and were removed from the final model.<br><br>*Depression symptoms:*<br>**Time (baseline > follow up)**<br><br>**Interaction time x isolation type**.<br>Post hoc analysis showed a **decrease of depression scores in the home group but not in the hotel group**.<br><br>*Anxiety symptoms:*<br>**Time (baseline > follow up)**<br><br>**Interaction time x isolation type**.<br>Post hoc analysis showed a **decrease of anxiety scores in the home group but not in the hotel group.** | Not reported |
| **Kopilas et al., 2021** [36] | Depression Anxiety Stress Scale-21 (DASS-21; depression and anxiety)<br><br>Impact of Event Scale-Revised (IES-R; distress subscales)<br><br>Positive and Negative Affect Schedule (PANAS; affect)<br><br>UCLA Loneliness Scale (ULS; loneliness) | One-way MANCOVA to examine effect of isolation type with post hoc tests for group differences.<br><br>*Isolation (exposed) compared to not in isolation (unexposed):*<br>Depression symptoms<br>Anxiety symptoms<br>Stress symptoms<br>Distress: Intrusion<br>**Distress: Hyperarousal (*isolation < not in isolation*)**<br>Distress: Avoidance<br>Positive affect<br>Negative affect<br>Loneliness | Not reported | Not reported |

| | | | | |
|---|---|---|---|---|
| | | **None of the group mean scores reached levels indicative of psychopathology* | | |
| **Kowalski et al., 2021** [37] | Psychological burden = reaching at least one of the cut-off scores of PHQ-8, GAD-7, or SSD-12:<br><br>Patient Health Questionnaire 8 (PHQ-8; depression).<br><br>Generalized Anxiety Disorder Scale 7 (GAD-7; anxiety).<br><br>Somatic Symptom Disorder-B Criteria Scale (SSD-12; distress). | Not reported | Mann-Whitney U-test (unadjusted),<br><br>*Psychological burden*<br>**Perceived stress (high > low)**<br>**Covid symptoms (high > low)**<br>**Poor physical health (high > low)**<br>**Perceived stressors (disgrace, job fear, social restrictions, infectiousness: (high > low)**<br><br>**Coping strategies (in general and in relation to COVID-19; low > high)**<br>**Social support (high > low)** | Not reported |
| **Maric et al., 2022** [39] | Mini International Neuropsychiatric Interview (MINI Standard 7.0.2; mood disorder, anxiety disorder, substance use disorder).<br><br>Patient Health Questionnaire-9 (PHQ-9; depression and anxiety symptoms).<br><br>General Anxiety Disorder-7 (GAD-7: anxiety symptoms)<br><br>Binarised using established cutoffs of symptom intensity | Logistic regression (odds ratios)<br><br>Any disorder<br>Mood disorder<br>Anxiety disorder<br>Substance use disorder | Not reported | Not reported |
| **Mohamed & Yousef, 2021** [40] | Arabic version of the Hospital Anxiety and Depression Scale (HADS; depression symptoms and anxiety syptoms)<br><br>Arabic version of the Davidson Trauma Scale (DTS; PTSD symptoms). | | Binary logistic regression (adjusted odds ratios)<br><br>*Anxiety symptoms*<br><br>***Depression symptoms* (home > hospital)**<br><br>***PTSD symptoms* (home < hospital)** | Not reported |
| **Navas et al., 2022** [41] | Impact of Events Scale – Revised (IES-R), adapted items (not reported) to create:<br><br>Frustration | Not reported | Binary logistic regression (adjusted odds ratio):<br><br>***Frustration* (home > hotel)**<br>***Anger/irritability* (home > hotel)** | Not reported |

| | | | | |
|---|---|---|---|---|
| | Anger/irritability<br>Feeling isolated<br>Anxiety<br><br>Rated on a scale of 0 to 5. The variables were coded as 'present' if it had been given a score of 2 to 5 (sometimes, often, almost always or always). | | | |
| **Oginni et al., 2021 [42]** | 14-item Hospital and Anxiety Scale (HADS; depression and anxiety subscales) | Multivariate linear regression<br><br>Depression symptoms (in both men and women; *isolation yes \|no*)<br>Anxiety symptoms (in both men and women; *isolation yes \|no*) | Multivariate linear regression<br><br>*Depression symptoms* (men \| women)<br>*Anxiety symptoms* (men \| women) | Not reported |
| **Opakunle, 2022 [43]** | Generalized Anxiety Disorder (GAD-7; anxiety symptoms)<br><br>Patient Health Questionnaire (PHQ-9; depression symptoms)<br><br>1 item from PHQ-9 was used to assess suicidal ideation<br><br>Insomnia Severity Index (ISI-7; insomnia)<br><br>Brief Self Rating Scale (BSRS-5; overall psychological symptoms)<br><br>All categorised using predetermined thresholds | Not reported | Binary logistic regressions (adjusted odds ratios):<br><br>***Psychological symptoms* (high viral load > low viral load)**<br>***Suicidal ideation* (high viral load > low viral load)**<br>*Anxiety*<br>*Depression*<br>*Insomnia* | |
| **Paz et al., 2020 [44]** | Patient Health Questionnaire (PHQ-9; depression symptoms)<br><br>Generalized Anxiety Disorder (GAD-7; anxiety symptoms)<br><br>Binarised using established cutoff scores indicating severity | Not reported | Logistic regression (adjusted odds)<br><br>*Depression*:<br>**Gender (female > male)**<br>**Region (coastal > other)**<br>**Regular schedule (no > yes)**<br>**Regular exercise (no > yes)**<br>**Time spent on COVID-19 information (1h < none; 1 < more than 1 h)** | Not reported |

| | | | | |
|---|---|---|---|---|
| | | | *Anxiety:*<br>**Gender (female > men)**<br>Region<br>**Regular schedule (no > yes)**<br>Regular exercise<br>**Time spent on COVID-19 information (1h < none; 1 < more than 1 h)** | |
| **Petrocchi et al., 2021** [45] | The Italian version of the NCCN Distress Thermometer (distress)<br><br>The Generalized Anxiety Disorder 7-item Scale (GAD-7; anxiety symptoms)<br>The Patient Health Questionnaire-9 (PHQ-9; depression symptoms).<br><br>For GAD-7 and PHQ-9, participants indicated how often they had been troubled during lockdown by each symptom, using a four-point Likert scale ranging from 0 ("Not at all") to 3 ("Nearly every day"). | Not reported | ANOVAs (separate unadjusted models for each outcome) and post hoc Duncan tests for group comparisons<br><br>*Depression* **(COVID pos home > COVID neg home > COVID neg home)**<br><br>*Anxiety* **(COVID pos hospital > COVID neg home)**<br><br>*Distress* (**COVID pos hospital > COVID neg home**) | Not reported |
| **Pheh et al., 2020** [46] | Subjective Unit of Distress Scale (SUDS; distress)<br><br>The Generalized Anxiety Disorder 7-item Scale (GAD-7; anxiety symptoms)<br><br>World Health Organization-Five Well-Being Index (WHO-5; general wellbeing) | MANCOVA to examine effect of self-isolation measured at baseline (yes/no) on psychological wellbeing outcomes at follow-up<br><br>Distress<br>**Anxiety (self-isolation > no self-isolation)**<br>General wellbeing | Not reported | Not reported*<br><br>**did not meet inclusion criteria for this review* |
| **Plesea-Condratovici et al., 2022** [47] | The Hospital Anxiety and Depression Scale (HADS; anxiety symptoms) | Not reported | Spearman's correlations (unadjusted)<br><br>*Anxiety*<br>**Gender (female > male)**<br>**Rural setting (yes < no)**<br>**Garden (yes < no)**<br>**Preexisting diagnosis of anxiety (yes > no)**<br>**No pulse oximeter (yes > no)** | Not reported |

| | | | | |
|---|---|---|---|---|
| **Rajagopalan et al., 2022** [48] | Depression Anxiety and Stress Scale-21 (DASS-21; total symptom score, depression score, anxiety score, stress score)<br><br>Pittsburgh Sleep Quality Index (PSQI; sleep quality)<br><br>WHO Quality of Life (WHOQOL-BREF; psychological subscale) | Not reported | Not reported | Paired t-test (unadjusted)<br><br>**Total symptoms (baseline > follow up)**<br>**Depression (baseline > follow up)**<br>Anxiety<br>Stress<br>**Insomnia (follow up > baseline)**<br>**Quality of life (follow up > baseline)** |
| **Ripon et al., 2020** [49] | Impact of Event Scale-Revised (IES-R; PTSD scores ≥20)<br><br>Center for Epidemiologic Studies–Depression scale (CES-D; Depression diagnosis scores ≥16) | Not reported | Binary logistic regressions (adjusted odds ratios)<br><br>*Depression*<br><br>***PTSD* (institutional > home)** | Not reported |
| **Rossi et al., 2020** [50] | The Global Psychotrauma Screen, post-traumatic stress symptoms subscale (GPS-PTSS; PTSS = 3/5)<br><br>The 9-item Patient Health Questionnaire (PHQ-9; severe depression symptoms ≥15)<br><br>The 7-item Generalized Anxiety Disorder scale (GAD-7; severe anxiety symptoms ≥15)<br><br>The 7-item Insomnia Severity Index (ISI; severe insomnia ≥22)<br><br>The 10-item Perceived Stress Scale (PSS; stress = higher quartile)<br><br>The International Adjustment Disorder Questionnaire (IADQ; adjustment | Binary logistic regression (adjusted odds ratios)<br><br>**PTSS (*currently in quarantine > not in quarantine*)**<br>Depression<br>**Anxiety (*currently in quarantine > not in quarantine*)**<br>Insomnia<br>Stress<br>**Adjustment disorder symptoms (*currently in quarantine > not in quarantine*)** | Not reported | Not reported |

| | | | | |
|---|---|---|---|---|
| | disorder symptoms = if associated with COVID-19) | | | |
| **Schluter et al., 2022** [51] | Generalized anxiety disorder (GAD-7; anxiety disorder ≥10)<br><br>Patient Health Questionnaire-9 (PHQ-9; major depressive episode ≥10) | Multivariate multilevel mixed-effects Poisson regression model (adjusted relative risk). No isolation as reference category.<br><br>**Probable anxiety disorder and/or depression disorder (*each quarantine/isolation group (contact, symptoms, diagnosis) > no isolation)*** | Pairwise comparisons (adjusted)<br><br>*Probable GAD and/or MDE* no significant difference identified between the isolation groups (contact, symptoms, diagnosis, travel) | Not reported |
| **Wessely et al., 2022** [53] | Wellbeing outcome: Alcohol use disorder identification test-consumption (AU-DIT-C)<br><br>Predictors:<br>Psychological burden – 5 items taken from the COVID-19 Snapshot Monitoring (COSMO) study<br><br>Coping strategies – 6 items taken from the COSMO study | Not reported | Logistic regression. (adjusted odds ratios)<br><br>*Increased alcohol consumption since before the pandemic*<br>**Isolation group (contacts > infected)**<br>**In partnership (yes > no)**<br>**At risk drinking behaviour (yes > no)**<br>**Coping strategies (no > yes)**<br>Age<br>Gender<br>Education<br>Psychological burden | Not reported |
| **Xu et al., 2020** [54] | Wellbeing outcome:<br>Zung's Self-Rating Anxiety Scale (SAS; anxiety symptoms)<br><br>Predictors:<br>The Chinese version of the Perceived Stress Scale (CPSS-14; stress, not reported how binarised)<br><br>The Cognitive Reappraisal subscale of the Emotion Regulation Questionnaire (EAIM; cognitive reappraisal, not reported how binarised) | Not reported | Moderation analysis using multivariate linear regression models.<br><br>*Anxiety*<br>**Stress (direct effect, positive association)**<br>**Cognitive reappraisal (direct effect, negative association)**<br>**Interaction (cognitive reappraisal moderates the association between stress and *anxiety*)** | Not reported |

*Table 3. Qualitative outcomes by research aim*

| Citation | Measures | Method of analysis | Impact of self-isolation on wellbeing | Factors perceived to be associated with wellbeing | Other potentially relevant data |
|---|---|---|---|---|---|
| **Domenghino et al., 2022** [26] | Questionnaire sections included medical history, details of SARS-CoV-2 (e.g. reason for testing, symptoms, severity), mental health prior to infection, isolation experience, and current physical and mental health; each section included a 'free text' space for participants to describe additional issues not captured by the survey responses | Comments concerning isolation or mental health were assigned to preliminary categories: circumstances of isolation, positive and negative aspects, mental health burden, and 'various based on the quantitative themes of the analysis'; specific words which came up frequently were also quantified through text search; the authors report 'themes that were repeatedly mentioned or that were considered especially impactful or important from a public health perspective' | Participants reported feelings of depression, loneliness, and aggression due to isolation, as well as feeling sad and that their mental health had worsened. Over '23 comments' (unclear whether this was 23 different people, or multiple comments from the same people) described isolation as being like a prison, torture, solitary confinement, or punishment<br><br>Feared stigma and workplace consequences<br><br>Relationships suffered: e.g. some participants reported conflicts with partners if their views on social contact differed, or feeling more aggressive due to isolation | Those already struggling with mental health pre-isolation felt that it worsened their symptoms<br><br>Worried about others they may have infected, especially their families<br><br>Perceived to be easier to cope with isolation if managers at work were understanding (vs. those who felt their managers 'blamed' them for getting sick or insisting they worked while sick, which added to their stress); managers calling frequently to check how they were doing also helped participants feel more positive<br><br>Not knowing the consequences of infection caused feelings of fear, panic, wondering if they would die<br><br>Isolation described as draining, tiring and discouraging, with participants describing somatic pain due to lack of physical activity and social contact; the more time they spent in isolation, the worse they felt physically<br><br>Participants who worked in health care were particularly concerned about 'abandoning' their colleagues | Phone calls and WhatsApp helped for a while but were no substitute for real contact<br><br>Some used the time to organise their homes, take up a new hobby, learn a language, read books, listen to podcasts, do yoga and home workouts |

| Citation | Measures | Method of analysis | Impact of self-isolation on wellbeing | Factors perceived to be associated with wellbeing | Other potentially relevant data |
|---|---|---|---|---|---|
| | | | | at a time of need, and did not feel protected in the workplace<br><br>Worries about financial troubles and job security were not frequent, but for those who experienced such worries, they were highly stressed<br><br>Those with children reported that the most stressful parts of isolation were being separated from children and the conflict between home office and childcare<br><br>Participants felt especially 'dissatisfied and alone' if they received contrary isolation instructions from different authorities<br><br>Positive aspects: more time with family, more time to relax, the ability to 'slow down' all helped participants to refocus on important things in life and appreciate what they had | |
| **Gok, 2022** [28] | Study-specific qualitative survey with 4 questions: how did you feel when you learned you were diagnosed with COVID-19; how did staying at home [in] quarantine affect you when you | Content analysis | 115 females and 39 males described psychological effects. Most frequently used codes in the qualitative descriptions of how participants felt about home quarantine: 'negative' (31.3% of 115 females, 23.1% of 39 males); 'loneliness' (13% of females, 15.4% of males); 'boring' (20% of females, | N/A | Aspects of quarantine noted to be particularly difficult:<br><br>35 females and 19 males discussed fears for their family, namely fear of their children being orphaned (51.5% of females, 68.5% of males) and loss |

| Citation | Measures | Method of analysis | Impact of self-isolation on wellbeing | Factors perceived to be associated with wellbeing | Other potentially relevant data |
|---|---|---|---|---|---|
| | were diagnosed with COVID-19; when you are diagnosed with COVID-19, can you describe the worst-case scenario you thought of while in quarantine; how did you cope with the uncertainty you were in when you were diagnosed with COVID-19? | | 10.2% of males); 'disturbing' (16.5% of females, 15.4% of males). Other codes included 'stressful', 'like a prison', 'despair' and 'sad'<br><br>14 females and 5 males discussed 'physiological effects', with home quarantine described as 'relaxing' by 64.3% of the females and 0 males, 'exhausting' by 35.7% of females and 60% of the 5 males, and 'difficult' by 0 females and 40% of the males.<br><br>The author also discusses the code 'ineffective' which was discussed by 23 females and 12 males. Home quarantine 'did not affect' 95.7% of these females and 8.3% of these males, and was described as 'mixed' by 4.3% of the females and 91.7% of the males – it is not clear what this means | | of family members (48.5% of females and 31.5% of males)<br><br>110 females and 33 males discussed physiological effects being the 'worst case scenario' they thought about during home quarantine, namely 'death' (31% of females, 21.2% of males), 'shortness of breath' (16.3% of females, 24% of males), 'being admitted to intensive care' (14.5% of females, 12% of males), 'transmitting the virus to others' (15.5% of females, 18% of males), 'serious damage to the body' (17.2% of females, 18% of males) and 'getting sick while in quarantine' (5.4% of females, 6% of males)<br><br>Codes emerging which related to coping with uncertainty included 'positive perspective' (encompassing 'positive thinking', acceptance, and 'trying to be strong' – 57 females and 28 males); 'maintaining a daily routine' (encompassing cleaning/watching movies/reading and healthy eating - 22 females and 9 males); 'social support' (encompassing communication with family/friends, partner support, |

| Citation | Measures | Method of analysis | Impact of self-isolation on wellbeing | Factors perceived to be associated with wellbeing | Other potentially relevant data |
|---|---|---|---|---|---|
| | | | | | 'getting strength from the presence of children' and getting information from healthcare professionals – 30 females and 12 males); and 'spirituality' (encompassing prayer, taking refuge in God and being patient – 19 females and 4 males) |
| **Jesmi et al., 2021** [33] | Semi-structured interviews including questions such as: describe your experience of COVID-19 infection; describe your feelings when informed of your test result; compare your COVID-19 experience with what others and the media describe; describe the experience of a day dealing with the disease; what comes to mind when you hear 'COVID-19 infection'?; what were your expectations from those around you?; 'what comes to mind when you think of a problem?' | Colaizzi phenomenological approach | Mental strains described by participants were divided into 3 categories: concerns, fears and isolation<br><br>Main fears were of death and disability/dependence on others<br><br>Most participants reported experiencing loneliness and boredom while quarantined at home | Concern about job loss was reported more frequently by males, self-employed participants and those with no job security<br><br>Daily media reports of COVID-19 deaths were perceived to contribute to fears<br><br>Experiencing shortness of breath was perceived to contribute to fear<br><br>Most participants reported similar concerns: worrying about worsening of symptoms (especially shortness of breath), losing their jobs, future of their children (e.g. what would happen to their children if their parent died), persisting complications, and disease disclosure (fear of telling families and causing them to be worried). Concerns were reported to lead to anxiety, tension, and sleep disorders (insomnia and nightmares) | Coping strategies included: religious activities / beliefs; complementary therapies such as vitamins, soup and herbal tea; relying on family support (including both mental and physical care) |

| Citation | Measures | Method of analysis | Impact of self-isolation on wellbeing | Factors perceived to be associated with wellbeing | Other potentially relevant data |
|---|---|---|---|---|---|
| **Lohiniva et al., 2021** [38] | Interviews including questions such as: describe any negative experiences with people during quarantine/isolation or any experiences of stigma; 'tell me about your time in quarantine'; how did quarantine influence your life?; what was difficult for you during quarantine? | Thematic analysis; data on stigma were analysed using a health stigma framework which entailed identifying codes and categories within each construct of the framework (drivers, facilitators, manifestations, outcomes, and impacts of stigma) whereas data on quarantine experiences were analysed based on an inductive coding process | Households with mild COVID-19 symptoms tended to describe quarantine as boring and monotonous<br><br>Some respondents described quarantine as having created tension at home whilst others saw positive developments, such as getting closer to their partner, having more family time, or having more time to relax | Stigma and self-stigma were perceived to be related to poorer quality of life: see next column<br><br>Households with severe or prolonged symptoms, or where household members had difficulty accessing testing or being admitted to hospital, tended to describe their quarantine experience as revolving around symptom management, worries about health and fears of death<br><br>For those isolating due to sick family members, caring for sick household members was considered a heavy responsibility consuming all their time and energy<br><br>Concerns were reported about health, symptoms, death<br><br>Some respondents with children worried their children were being ignored while they managed the illness among other household members<br><br>Knowing someone hospitalised due to COVID-19 and seeing media reports about high infection rates and fatalities abroad were perceived to worsen thoughts/worries about death (many described media | Stigma:<br><br>Facilitators of stigma: fear of contracting coronavirus, blame for contracting coronavirus<br><br>Manifestations of stigma: blame for being irresponsible, gossiping, being overly curious about their COVID-19 experience, reluctance to interact<br><br>Stigma outcomes: fear of disclosing COVID-19 status<br><br>Stigma impact: negative influence on social contacts resulting in reduced quality of life<br><br>Self-stigma:<br><br>Facilitators: different and conflicting information about COVID-19, symptoms continuing after quarantine, uncertainty about immunity, uncertainty about having contracted COVID-19<br><br>Manifestations: stressed when outside the home, nervous to meet people, fear of being blamed for leaving the house<br><br>Outcomes: reluctance to meet people, prolonging quarantine/isolation periods |

| Citation | Measures | Method of analysis | Impact of self-isolation on wellbeing | Factors perceived to be associated with wellbeing | Other potentially relevant data |
|---|---|---|---|---|---|
| | | | | coverage as exhausting and stressful but others felt it was helpful and allowed them to prepare themselves for having COVID-19)<br><br>Respondents with active and outgoing lifestyles pre-quarantine were 'particularly bothered' by the restrictive quarantine (vs. those who had little social contact outside of the home and/or worked from home pre-quarantine, who did not feel their lives had changed very much as a result of quarantine)<br><br>Worries about the health of family members and guilt at feeling they had infected family members | Impacts: psychological distress leading to reduced quality of life<br><br>Other quarantine experiences:<br><br>Peer support groups (e.g. WhatsApp groups for people who attended an event where they contracted the virus, or group chats with friends who also had COVID-19) were seen by some participants as very valuable and empowering and they found it helpful to talk to others going through similar experiences, who understood them; some also said this communication helped reduce their worries about their own health and that of their families; such groups were also seen as a good source of information; however, other participants felt that continuous discussions about the illness in these groups made the illness highly present in their lives and a source of stress |
| **Verberk et al., 2021** [52] | Semi-structured interviews covering the following topics: experience of home care, transmission prevention practices, impact (of household | Researchers summarised data directly from the audio recordings using an a priori framework that captured key areas of interest based on the research questions | Emotional burden: participants described living with and caring for a relative with COVID-19 as having a significant emotional impact. Some described feelings of helplessness and lack of control | Participants expressed concerns about having no clear instructions on how to monitor their infected loved ones; the potential for symptoms to suddenly worsen; and practicalities such as finding | Stigma: participants described fear of reactions of others who were scared of being infected themselves; they felt judged and seen as contagious and felt people were afraid to be near them |

| Citation | Measures | Method of analysis | Impact of self-isolation on wellbeing | Factors perceived to be associated with wellbeing | Other potentially relevant data |
|---|---|---|---|---|---|
| | members' COVID-19 diagnosis) on daily life | | whilst others described acceptance and 'taking it day by day'<br><br>Some worried about catching the virus (and potential health impacts of that) and transmitting the virus to others<br><br>Many had experienced an initial reaction of anger towards the infected person; however, participants also expressed feelings of solidarity within the household<br><br>Participants described feelings of boredom, lack of control and feeling 'depressed' | childcare if they needed to go to hospital<br><br>Participants perceived that it was more difficult to be the non-infected person in the household because those who had been infected could return to work after symptoms had been resolved for at least 14 days, whereas contacts often found their quarantine duration extended as different household members contracted the virus and some had to quarantine for two months<br><br>Participants described confusion over inconsistent guidelines and felt that rules were not communicated efficiently. Multiple sources of differing information could create information overload and confusion<br><br>Quarantine was frustrating because they had no clear idea of how long they would need to quarantine for, and some were frustrated at having to enter quarantine just as lockdown measures relaxed | Participants stressed the importance of staying positive and accepting the situation. Coping strategies included talking to family members and wider support networks, finding time to relax, and making plans for post-quarantine |

### Appendix 7: Risk of bias summaries

1. *Quality appraisal for Aim 1 using ROBINS-E for exposure studies, summary by study (left) and domains (right)*

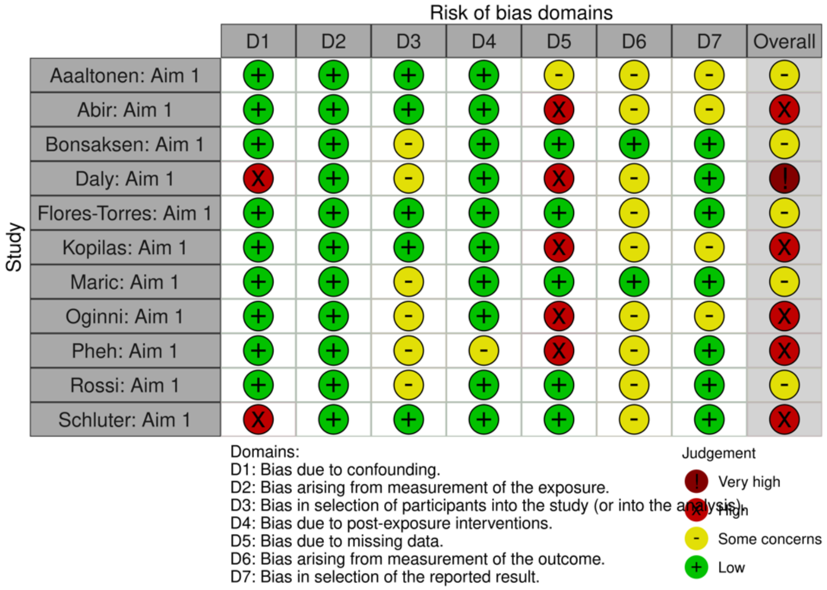

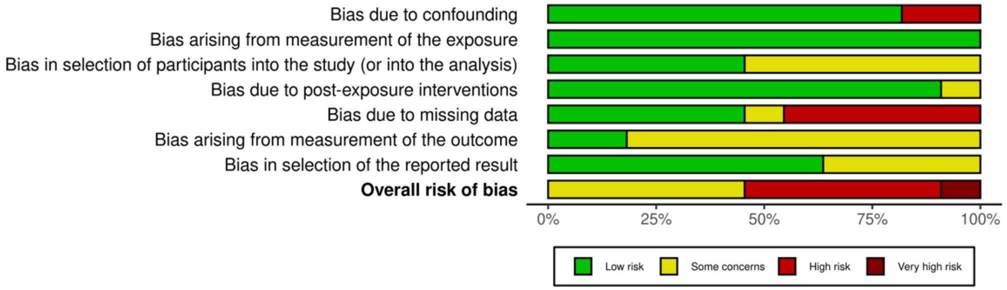

Note. Data visualisation created using McGuinness, L. A., & Higgins, J. P. T. (2020). Risk-of-bias VISualization (robvis): An R package and Shiny web app for visualizing risk-of-bias assessments. *Research Synthesis Methods*. https://doi.org/10.1002/jrsm.1411

2. *Quality appraisal for Aim 2 using ROBINS-E for exposure studies*

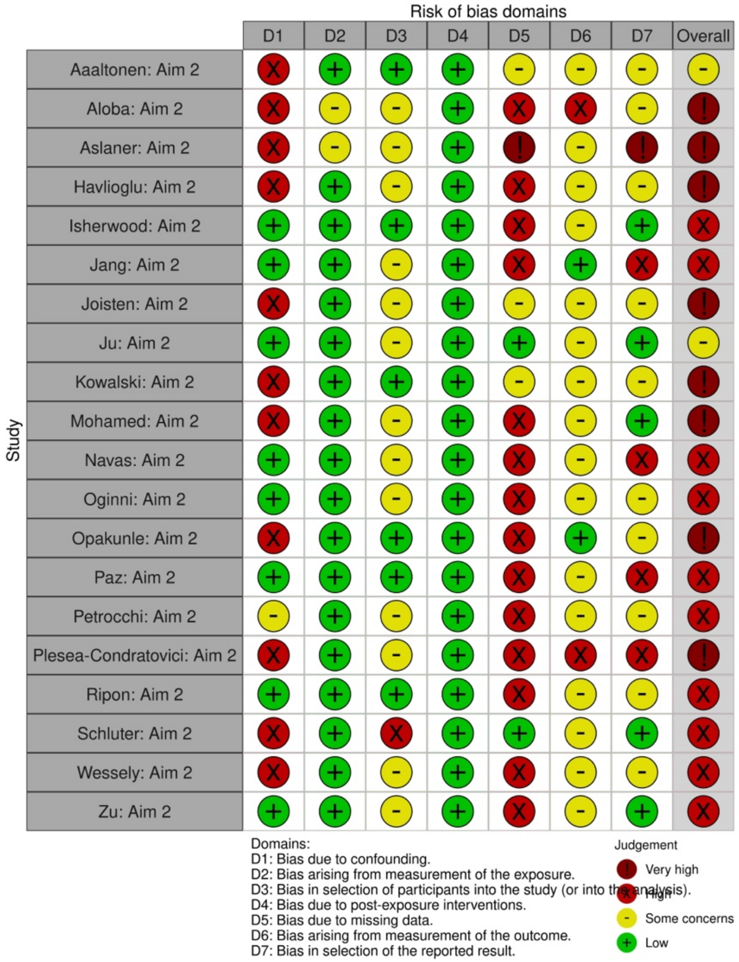

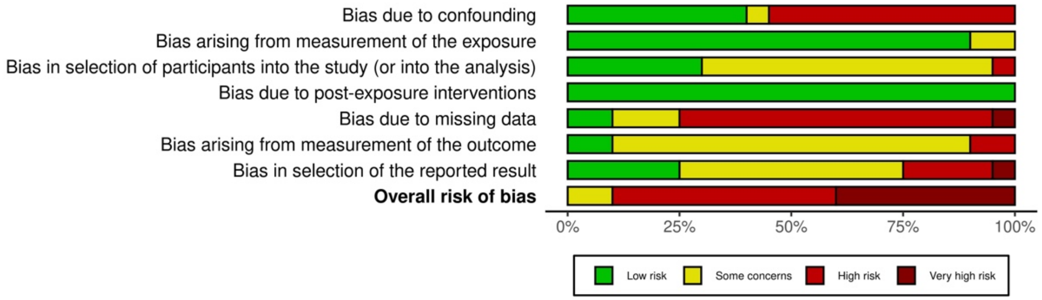

Note. Data visualisation created using McGuinness, L. A., & Higgins, J. P. T. (2020). Risk-of-bias VISualization (robvis): An R package and Shiny web app for visualizing risk-of-bias assessments. *Research Synthesis Methods*. https://doi.org/10.1002/jrsm.1411

*3. Quality appraisal for Aim 2 using ROBINS-I for non-randomised intervention studies*

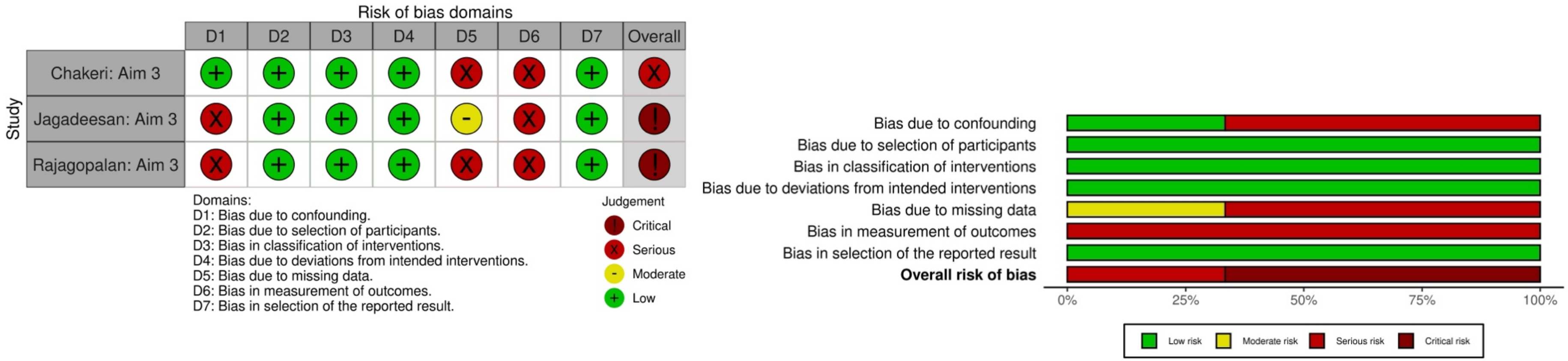

Note. Data visualisation created using McGuinness, L. A., & Higgins, J. P. T. (2020). Risk-of-bias VISualization (robvis): An R package and Shiny web app for visualizing risk-of-bias assessments. *Research Synthesis Methods*. https://doi.org/10.1002/jrsm.1411

*4. Quality appraisal – CASP checklist for qualitative studies*

| **Citation** | **Q1** | **Q2** | **Q3** | **Q4** | **Q5** | **Q6** | **Q7** | **Q8** | **Q9** | **Q10** | **% of 'yes' answers** | **Other comments** |
|---|---|---|---|---|---|---|---|---|---|---|---|---|
| Domenghino et al., 2022 [26] | Yes | Yes | No | No | No | No | Yes | No | No | Yes | 40% | - Analysis not described well and does not appear to be rigorous<br>- Data were categorised into pre-defined categories according to what the authors expected to find / found important<br>- Not reported how many qualitative comments there were or from how many people<br>- Participants who did not meet the study's inclusion criteria were included (e.g. lowest age was 17, but 17-year-olds should have been excluded) |
| Gok, 2022 [28] | Yes | Can't tell | No | No | No | No | Yes | No | No | No | 20% | - Population size is reported differently in different places in the paper (202 vs 212)<br>- Date of data collection is reported differently in different places in the paper<br>- Wording of Q4 on the survey is leading<br>- Codes are not well-explained<br>- Quotes presented do not always relate well to the 'codes' they have been given<br>- Unclear why the %s always add up to 100% in the tables – could participants not give more than one answer? This is not explained |

| | | | | | | | | | | | | |
|---|---|---|---|---|---|---|---|---|---|---|---|---|
| | | | | | | | | | | | | - Author describes rates being 'higher in females' but they do not perform any statistical analysis to know whether this is a significant finding<br>- Very little discussion of implications of the study |
| Jesmi et al., 2021 [33] | Can't tell | Can't tell | Can't tell | Can't tell | Yes | No | No | No | Yes | No | 20% | - Aim is very vague ('lived experiences' of those with COVID-19)<br>- Ethical approval was granted but only 8/14 participants completed informed consent forms |
| Lohiniva et al., 2021 [38] | Yes | Yes | Yes | No | No | No | Yes | No | Yes | Yes | 60% | |
| Verberk et al., 2021 [52] | Yes | Yes | Yes | Can't tell | Yes | No | Yes | No | Yes | Yes | 70% | - Explanation of how analysis was done is insufficient; potential for bias due to the a priori framework based on 'areas of interest' |

**Key:**

- **Q1: Was there a clear statement of the aims of the research?**
- **Q2: Is a qualitative methodology appropriate?**
- **Q3: Was the research design appropriate to address the aims of the research?**
- **Q4: Was the recruitment strategy appropriate to the aims of the research?**
- **Q5: Was the data collected in a way that addressed the research issue?**
- **Q6: Has the relationship between researcher and participants been adequately considered?**
- **Q7: Have ethical issues been taken into consideration?**
- **Q8: Was the data analysis sufficiently rigorous?**
- **Q9: Is there a clear statement of findings?**
- **Q10: Do the authors discuss the value of the research in terms of implications and contributions to literature? (please note Q10 has been reworded from 'How valuable is the research?' to enable yes/no**

**Appendix 8: Full narrative description of factors associated with psychological wellbeing during self-isolation**

In the risk of bias assessments, no findings were at low risk, one had some concerns, [35] and most findings were at high, [21,30,32,41,42,44,45,49,51,53] or very high risk of bias. [19,22,29,34,37,40,43,47,54]; Appendix 7). The domains that were high risk overall were bias due to missing data (79%), confounding (53% of studies), selection of the reported result (26%), the outcome measurement (11%), and participant selection (5%). # Comparing findings between risk of bias ratings did not alter the interpretation of the findings for aim 2.

Factors related to self-isolation were reported in eleven studies. Factors included COVID-19 related stressors, [32,37,44] the reason for self-isolation, [19,34,45] access to support, [32,37] duration, [21,35] and location. [40,45,49,51] COVID-19 stressors and a lack of support during self-isolation were both consistently associated with higher levels of general, depressive, and anxiety symptoms. [32,37,44] Symptoms of depression and anxiety were found to be lower at the end of self-isolation compared to the start, [35] and a longer period of self-isolation was associated with lower sleep problems at the end of the isolation period. [21] The reason for self-isolation (infection or close contact) was not consistently reported to associate with general psychological symptoms, [19,34] or anxiety, [19,45] but both studies that examined depression reported that symptoms were higher when people isolated due to infection. [19,45] Self-isolation at home rather than a hotel was associated with lower PTSD symptoms, [40,49] but was not associated with a change in anxiety symptoms. [40,45,51] Evidence for a relationship between the location of self-isolation and depression was unclear, [40,45,49,51] although the outcomes used to measure depression were robust and there were no other obvious differences between the studies that might explain the inconsistent findings.

Factors related to demographics were reported in six studies. Factors included living alone, [30,32] age, [21,30,32] and gender. [30,32,42,44,47] Living alone was not found to associate with general or depression symptoms. [30,32] However, there was no consistent evidence for an association between age or gender with any wellbeing outcome. [30,32,42,44,47]

Factors related to mental or physical health were reported in five studies, all of which reported a consistent negative impact on wellbeing outcomes. Psychological disorders and symptoms before or during self-isolation were associated with psychological burden, [37] anxiety symptoms, [47,54] and insomnia. [21] Poor physical health was associated with psychological burden, [37] and depression symptoms. [32] Low levels of coping strategies were associated with psychological burden, [37] and anxiety symptoms. [54]

Factors related to COVID-19 were reported in three studies, including higher viral load and more severe symptoms. [21,37,43] Both these factors were associated with an increase in general psychological symptoms. [37,43] Whereas no evidence was found for an association with depression, anxiety, or insomnia. [21,43]

Qualitative findings largely supported the factors identified in the quantitative findings (Table 3). For example, participants perceived that pre-existing mental health problems worsened during self-isolation. [26] Fears around COVID-19 were also prominent and appeared to contribute to poor wellbeing. Participants described concerns about the consequences of infection, [26] the experience of COVID-19 symptoms, [33,38] fears of death or dependence, [33] concerns about workplace consequences and stigma, [26] worries about others due to knowledge of high infection rates and fatalities, [38] and fears of transmitting the virus to others. [26,33,38,52] These concerns were perceived to lead to anxiety and sleep problems (43). Other factors which participants perceived to increase the negative psychological impacts of self-isolation included financial difficulties, [26,33] stigma and self-stigma, [38,52] and exposure to excessive media coverage of COVID-19 and conflicting guidance. [26,33,38,52] In addition, some people also reported a positive impact on their wellbeing, such as having more time to spend with family and to relax, which allowed them to refocus and appreciate what they had. [26,38,52]. Notably, the studies rated as highest in quality were the only two which reported on stigma. [38,52](Appendix, 7) Concerns about COVID-19 symptoms, fears of transmitting the virus, and exposure to media coverage and conflicting guidance were reported as concerns in both lower- and higher-quality studies. Positive psychological effects were also reported in both lower- and higher-quality studies.

Importantly, qualitative findings indicated that the same factor could have a positive or negative impact on wellbeing, depending on the person's individual context. For example, having children was often reported as a risk factor for a negative impact on wellbeing during isolation, and one context under which this would occur was when there was a perceived conflict with childcare and work priorities. [26] On the other hand, some people who were worried during their isolation reported that the presence of children reduced stress and helped them to cope. [28] Uncertainties also seemed to play an important role that changed depending on context. For example, for those who were infected, not knowing how bad symptoms might get impacted their wellbeing, [26] whereas for those who were isolating due to infection within their household, not knowing how long isolation might go on for impacted their wellbeing. [33,38]
